# Emergence and evolution of *Plasmodium falciparum* histidine-rich protein 2 and 3 deletion mutant parasites in Ethiopia

**DOI:** 10.1101/2021.01.26.21250503

**Authors:** Sindew M. Feleke, Emily N. Reichert, Hussein Mohammed, Bokretsion G. Brhane, Kalkidan Mekete, Hassen Mamo, Beyene Petros, Hiwot Solomon, Ebba Abate, Chris Hennelly, Madeline Denton, Corinna Keeler, Nicholas J. Hathaway, Jonathan J. Juliano, Jeffrey A. Bailey, Eric Rogier, Jane Cunningham, Ozkan Aydemir, Jonathan B. Parr

**Author notes:** Co-first authors (Sindew M. Feleke, Emily N. Reichert). Co-senior authors (Jane Cunningham, Ozkan Aydemir, Jonathan B. Parr).

## Abstract

Malaria diagnostic testing in Africa is threatened by *Plasmodium falciparum* parasites lacking histidine-rich protein 2 (*pfhrp2*) and 3 (*pfhrp3*) genes. Among 12,572 subjects enrolled along Ethiopia’s borders with Eritrea, Sudan, and South Sudan and using multiple assays, we estimate HRP2-based rapid diagnostic tests would miss 9.7% (95% CI 8.5-11.1) of falciparum malaria cases due to *pfhrp2* deletion. Established and novel genomic tools reveal distinct subtelomeric deletion patterns, well-established *pfhrp3* deletions, and recent expansion of *pfhrp2* deletion. Current diagnostic strategies need to be urgently reconsidered in Ethiopia, and expanded surveillance is needed throughout the Horn of Africa.

---

*Plasmodium falciparum* strains that evade diagnosis by rapid diagnostic tests (RDTs) represent a major threat to malaria control and elimination efforts^1,2^. Malaria RDTs detect antigens produced by *Plasmodium* parasites, including *P. falciparum* histidine-rich protein 2 (HRP2), parasite lactate dehydrogenase (LDH), and aldolase. HRP2 has advantages over other biomarkers due to its abundance in the bloodstream, repetitive binding epitopes, and falciparum-specificity^3,4^. Most HRP2-based RDTs also exhibit some cross-reactivity to a closely related protein (HRP3). HRP2-based RDTs are currently the predominant malaria diagnostic test employed throughout sub-Saharan Africa^5–7^.

Deletion mutations involving the histidine-rich protein 2 and/or 3 (*pfhrp2/3*) genes allow parasite strains to escape HRP2-based RDT detection^8,9^. First described in clinical samples from Peru in 2010, these subtelomeric deletions on chromosomes 8 (*pfhrp2*) and 13 (*pfhrp3*) are frequently large (≥20kb), encompass multiple genes, and are difficult to study using existing methods^9–11^. Improved PCR and serological approaches can be used to increase confidence in deletion prevalence estimates^12–14^, but our understanding of the evolutionary history of *pfhrp2/3-*deleted *P. falciparum* is limited and largely informed by analysis of a small number of microsatellite markers^15–17^. Recent genomic analyses have begun to expand our understanding of *pfhrp2/3-*deleted *P. falciparum*^*18–20*^ but continue to be hindered by the challenges of assembling the highly repetitive and paralogous sequences of *P. falciparum*’s subtelomeres^21^. New tools are needed to support surveillance of *pfhrp2/3* deletions and to determine their true prevalence and the forces impacting their evolution and spread.

Increasing reports of these “diagnostic resistant” *pfhrp2/3-*deleted parasites in Africa in 2017-2018 prompted calls for urgent surveillance in affected regions, including countries in the Horn of Africa like Ethiopia^15,22–25^. Ethiopia is Africa’s second most populous country, and 68% of its population is at risk of malaria exposure^26^. *P. falciparum* infection accounts for the majority of malaria deaths^27^ and approximately 70% of all cases^26^. RDTs were first introduced in Ethiopia in 2004, and the country’s current test-treat-track strategy requires parasitological confirmation either by quality microscopy or RDT prior to antimalarial treatment^28^. *P. falciparum-Plasmodium vivax* (HRP2/Pv-specific-LDH) combination RDTs are the sole diagnostic test used in most settings. Over the last decade, Ethiopia has achieved remarkable progress in the fight against malaria through strong preventative and case management interventions, including engagement of volunteers to provide diagnostic services at a local level^28^. Reports of highly prevalent *pfhrp2/3-*deleted parasites in neighboring Eritrea suggest that these gains could be threatened^15,24^. Rapid assessment of the epidemiology of *pfhrp2/3* deletions in Ethiopia and surrounding regions is required to determine whether a change in malaria diagnostic testing policy is warranted.

Here, we describe the first prospective, multi-site study of *pfhrp2/3-*deleted *P. falciparum* in sub-Saharan Africa based on WHO’s *pfhrp2/3* deletion surveillance protocol^29^, released in 2018 to encourage a harmonized and representative approach to *pfhrp2/3* deletion surveillance and accurate reporting. Including sites spanning Ethiopia’s borders with Eritrea, Sudan, and South Sudan, we apply both established and novel genomic tools to determine the genetic epidemiology of *pfhrp2/3-*deleted *P. falciparum*, confirming deletions using multiple PCR assays^14^, an ultrasensitive bead-based immunoassay for antigen detection^13^, whole-genome sequencing (WGS)^30^, and/or molecular inversion probe (MIP) deep sequencing^31^. Using a MIP panel designed for high-throughput *pfhrp2/3* genotyping, we map and categorize deletion breakpoints and evaluate their flanking regions for evidence of recent evolutionary pressure favoring *pfhrp2/3-*deleted parasites in Ethiopia.

## RESULTS

### Study Population and RDT Results

A total of 12,572 study participants (56% male, 44% female) between the ages of 0 to 99 years who presented with one or more of symptoms consistent with malaria at 108 health facilities in the Amhara, Tigray, and Gambella regions between November 2017 and April 2018 were enrolled (**Table 1**). Median participant age was 19 years (interquartile range [IQR]: 8-30). From the same finger prick, participants were tested with two RDTs, including the routine HRP2/Pv-specific-LDH RDT combination test [CareStart *Pf/Pv* RDT (Access Bio, Somerset, NJ; product code RM VM-02571)] and the survey HRP2/Pf-specific-LDH RDT [SD Bioline Malaria Ag P.f. RDT (Alere, Waltham, MA; product code 05FK90)].

**Table 1.** Characteristics of study subjects and RDT results. Abbreviations: SD, standard deviation; RDT, rapid diagnostic test; HRP2, histidine-rich protein 2; Pv-LDH, *P. vivax* parasite lactate dehydrogenase; Pf-LDH, *P. falciparum* parasite lactate dehydrogenase.

|  | **Amhara** | **Gambella** | **Tigray** | **Overall** |
| --- | --- | --- | --- | --- |
| **Subjects, n** | 3,879 | 2,335 | 6,357 | 12,572 |
| **Age, median years (IQR)** | 20 (10-28) | 12 (5-19) | 21 (9-37) | 19 (8-30) |
| **Female sex, n (%)** | 1,492 (38.5) | 1,055 (45.2) | 3,008 (47.3) | 5,555 (44.2) |
| **Location, n (%)** |  |  |  |  |
| Rural | 2,350 (60.6) | 923 (39.5) | 4,445 (69.9) | 7,718 (61.4) |
| Urban | 1,282 (33.0) | 82 (3.5) | 1,609 (25.3) | 2,973 (23.6) |
| Missing | 247 (6.4) | 1,330 (57.0) | 303 (4.8) | 1,881 (15.0) |
| **Fever, n (%)** | 3,607 (93.0) | 2,255 (96.6) | 5,593 (88.0) | 11,455 (91.1) |
| **CareStart RDT, n (%)** |  |  |  |  |
| HRP2+, Pv-LDH+ | 59 (1.5) | 509 (21.8) | 25 (0.4) | 593 (4.7) |
| HRP2+ only | 1,053 (27.1) | 165 (7.1) | 507 (8.0) | 1,725 (13.7) |
| Pv-LDH+ only | 241 (6.2) | 11 (0.5) | 338 (5.3) | 590 (4.7) |
| Negative | 2,518 (64.9) | 1,650 (70.7) | 5,486 (86.3) | 9,654 (76.8) |
| Invalid | 8 (0.2) | 0 (0.0) | 1 (0.0) | 9 (0.1) |
| **SD-Bioline RDT, n (%)** |  |  |  |  |
| HRP2+, Pf-LDH+ | 719 (18.5) | 552 (23.6) | 276 (4.3) | 1,547 (12.3) |
| HRP2+ only | 297 (7.7) | 106 (4.5) | 201 (3.2) | 604 (4.8) |
| Pf-LDH+ only | 239 (6.2) | 11 (0.5) | 168 (2.6) | 418 (3.3) |
| Negative | 2,609 (67.3) | 1,665 (71.3) | 5,705 (89.7) | 9,979 (79.4) |
| Invalid | 15 (0.4) | 0 (0.0) | 2 (0.0) | 17 (0.1) |

Overall, 2,714 (22%) study participants were *P. falciparum* positive by at least one RDT (any HRP2 or Pf-LDH positive band); among these, 361 (13.3%; 95% confidence interval [CI] 12.1-14.7%) had a discordant RDT profile suggestive of *pfhrp2/3-*deleted *P. falciparum* infection, which was defined as HRP2-negative by both RDTs but Pf-LDH positive. Among the 2,714 samples that were *P. falciparum* positive by RDT, the northern region of Tigray had the highest proportion of infections with discordant RDT profiles at 140/689 (20.4%; 95% CI 17.5-23.7%), followed by Amhara with 211/1342 (15.8%; 13.9-17.8) and Gambella with 10/683 (1.5%; 0.7-2.8), as shown in **Figure 1**.

**Figure 1.**
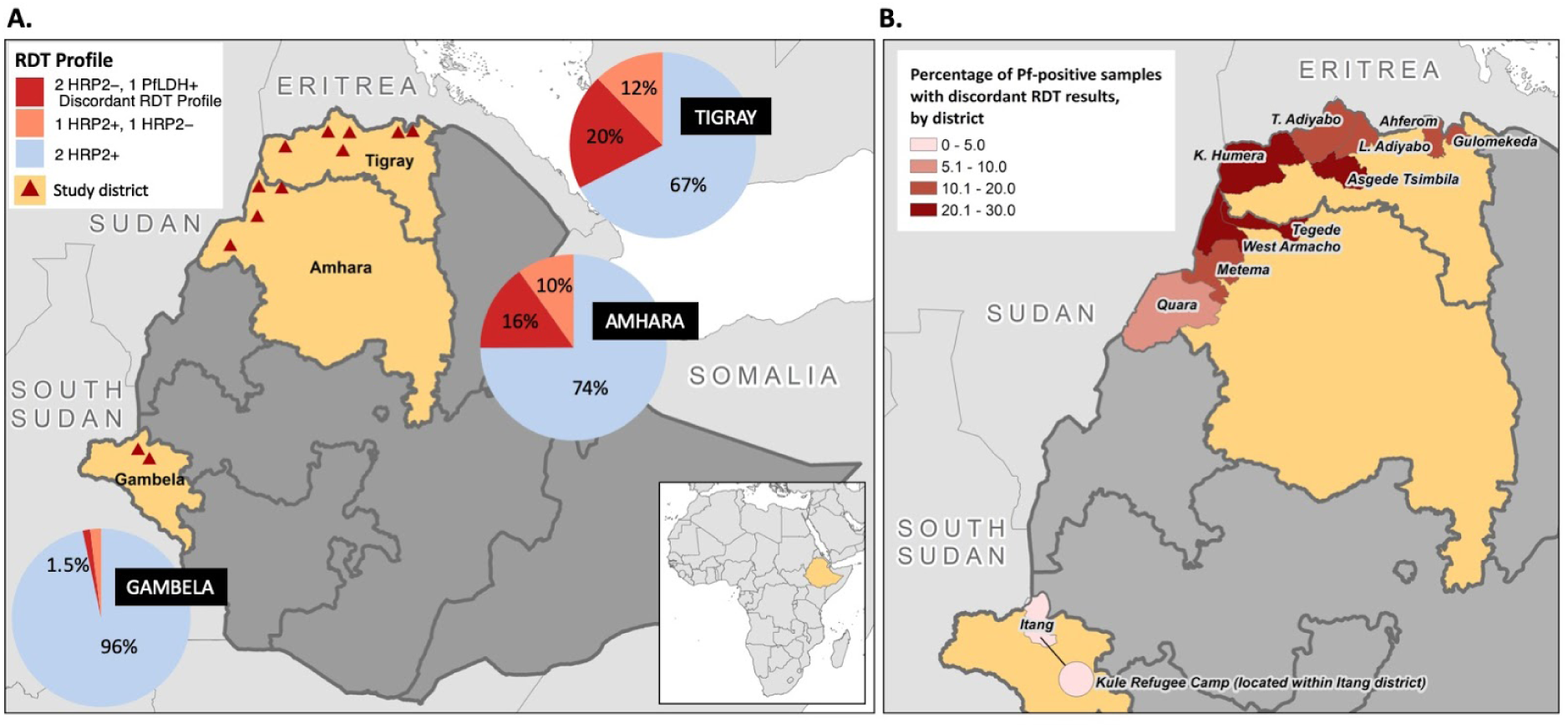
Distribution of P. falciparum-positive RDT results and discordant profiles suggestive of pfhrp2/3 gene deletions. A) Aggregated results from both RDTs, CareStart *Pf/Pv* (HRP2/Pv-LDH) RDT and SD Bioline Malaria Ag P.f. (HRP2/Pf-LDH) RDT, displayed by region for all *P. falciparum* infections. The ‘2 HRP2-, 1 Pf-LDH+’ discordant RDT profile indicates potential infection by *pfhrp2/3-*deleted *P. falciparum*. Triangles represent the enrollment sites, including 11 districts and the Kule refugee camp within the Itang district in Gambella. B) The percentage of study participants identified with *P. falciparum* infection by RDT who had the discordant RDT profile, by district.

### Pfhrp2/3 deletion PCR genotyping

820 samples with complete demographic and clinical data from Amhara (n = 524), Tigray (n = 225), and Gambella (n = 71) underwent molecular analysis. These samples were collected from subjects with the discordant RDT profile and a subset of subjects with other RDT results (**Supplementary Figure 1**), including a randomly selected 248/361 (68.7%) of those with the discordant RDT profile of interest (HRP2-, Pf-LDH+), as well as 465/2115 (22.0%) randomly selected *P. falciparum* RDT HRP2 positives (HRP2+, Pf-LDH+) as controls. The remaining 107 samples included 90 with inconclusive HRP2 results (HRP2+ by only 1 RDT, of which 67 were Pf-LDH- and 23 were Pf-LDH+) and 17 negative controls (HRP2-, Pf-LDH-). Quantitative real-time PCR (qPCR) targeting the *P. falciparum* lactate dehydrogenase (*pfldh*) gene confirmed parasitemia in 731/820 (89%) samples, with a geometric mean (GM) of 1,390.7 parasites/µL (geometric standard deviation [geoSD]: 9.8). Further analysis was restricted to the 610 samples with >100 parasites/µL to avoid misclassification of *pfhrp2/3* deletions due to low parasitemia (**Supplementary Figure 2**); 176 (28.9%) had the discordant RDT profile.

**Figure 2.**
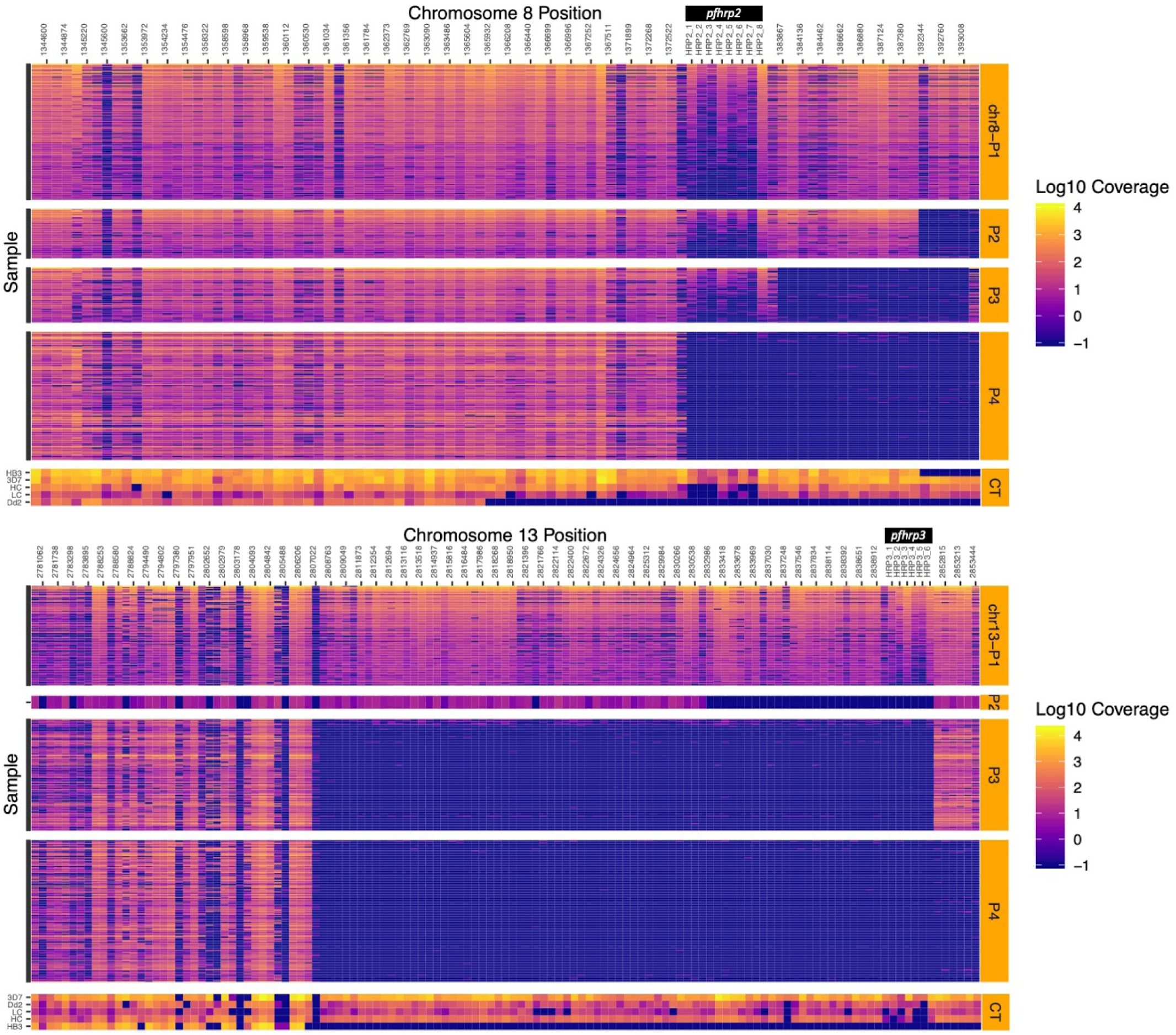
Deletion profiling using MIP sequencing of pfhrp2 (chromosome 8), pfhrp3 (chromosome 13), and flanking regions applied to 375 field samples. Samples are grouped by subtelomeric structural profile, with control strains denoted CT, as labeled along the right y-axis. Columns represent each MIP target segment, rows represent individual samples, and the color scale represents log_10_ unique molecular identifier (UMI) depth-of-coverage at each location. Columns are labeled by the midpoint of each probe’s target region.

Infection by *pfhrp2/3-*negative parasites was common among these 610 subjects when assessed by PCR, with 355 (58%; 95% CI 54-62) lacking detectable *pfhrp2* and/or *pfhrp3*, and 136 (22%; 19-26) lacking both *pfhrp2* and *pfhrp3*. For those lacking only one gene, *pfhrp3*-negative infections [192 (31%; 28-35) *pfhrp2+/pfhrp3*-] were more prevalent than *pfhrp2*-negative infections [27 (4.4%; 3-6) *pfhrp2*-/*pfhrp3+*]. Concordance between *pfhrp2-*negative PCR results and the discordant RDT profile was good (Cohen’s kappa 0.66). Overall, among samples with the discordant RDT profile, 64.8% (95% CI 57-72) were *pfhrp2-/3-* and 8.0% (5-13) *pfhrp2-/3+*, with an additional 15.9% (11-22) *pfhrp2+/3-* (**Supplementary Figure 3**). Interestingly, of samples HRP2+ by both RDTs, *pfhrp3* could not be amplified in 42.6% (38-48). We observed expected agreement between the results of *pfhrp2/3* PCR assays, RDTs, and a bead-based HRP2 immunoassay applied to a randomly selected subset of samples (see **Supplementary Results, Table 2**). No associations between *pfhrp2/3* PCR result and age, sex, or parasitemia were identified (**Supplementary Table 1**).

**Table 2.** Assay results across platforms. PCR, bead-based antigen immunoassay, molecular inversion probe (MIP) deep sequencing, and whole-genome sequencing (WGS) for samples *P. falciparum*-positive by RDT are shown.

|  | **RDT results** |  |  |
| --- | --- | --- | --- |
|  | 2 HRP2+, 1 PfLDH+ | 2 HRP2-, 1 PfLDH+ | 1 HRP2+, 1 HRP2- |
| **PCR result, n** | 379 | 176 | 47 |
| *pfhrp2*+/*3*+ | 210 (55%) | 20 (11%) | 19 (41%) |
| *pfhrp2*-/*3*- | 9 (2%) | 114 (65%) | 10 (22%) |
| *pfhrp2*-/*3*+ | 7 (2%) | 14 (8%) | 5 (11%) |
| *pfhrp2*+/*3*- | 152 (40%) | 28 (16%) | 12 (26%) |
| **HRP2 immunoassay result, n** | 243 | 167 | 42 |
| HRP2+ | 224 (92%) | 40 (24%) | 30 (71%) |
| HRP2- | 19 (8%) | 127 (76%) | 12 (29%) |
| **MIP result, n** | 198 | 104 | 30 |
| *pfhrp2*+/*3*+ | 77 (39%) | 5 (5%) | 5 (17%) |
| *pfhrp2*-/*3*- | 10 (5%) | 84 (81%) | 10 (33%) |
| *pfhrp2*-/*3*+ | 3 (2%) | 1 (1%) | 4 (13%) |
| *pfhrp2*+/*3*- | 108 (55%) | 14 (13%) | 11 (37%) |
| **WGS result, n\*** | 0 | 22 | 0 |
| *pfhrp2*+/*3*+ | NA | 0 (0%) | NA |
| *pfhrp2*-/*3*- | NA | 22 (100%) | NA |
| *pfhrp2*-/*3*+ | NA | 0 (0%) | NA |
| *pfhrp2*+/*3*- | NA | 0 (0%) | NA |

\* Zero median WGS coverage across 1,000 base-pair windows encompassing *pfhrp2* or *pfhrp3*, in samples with clinical data.

**Figure 3.**
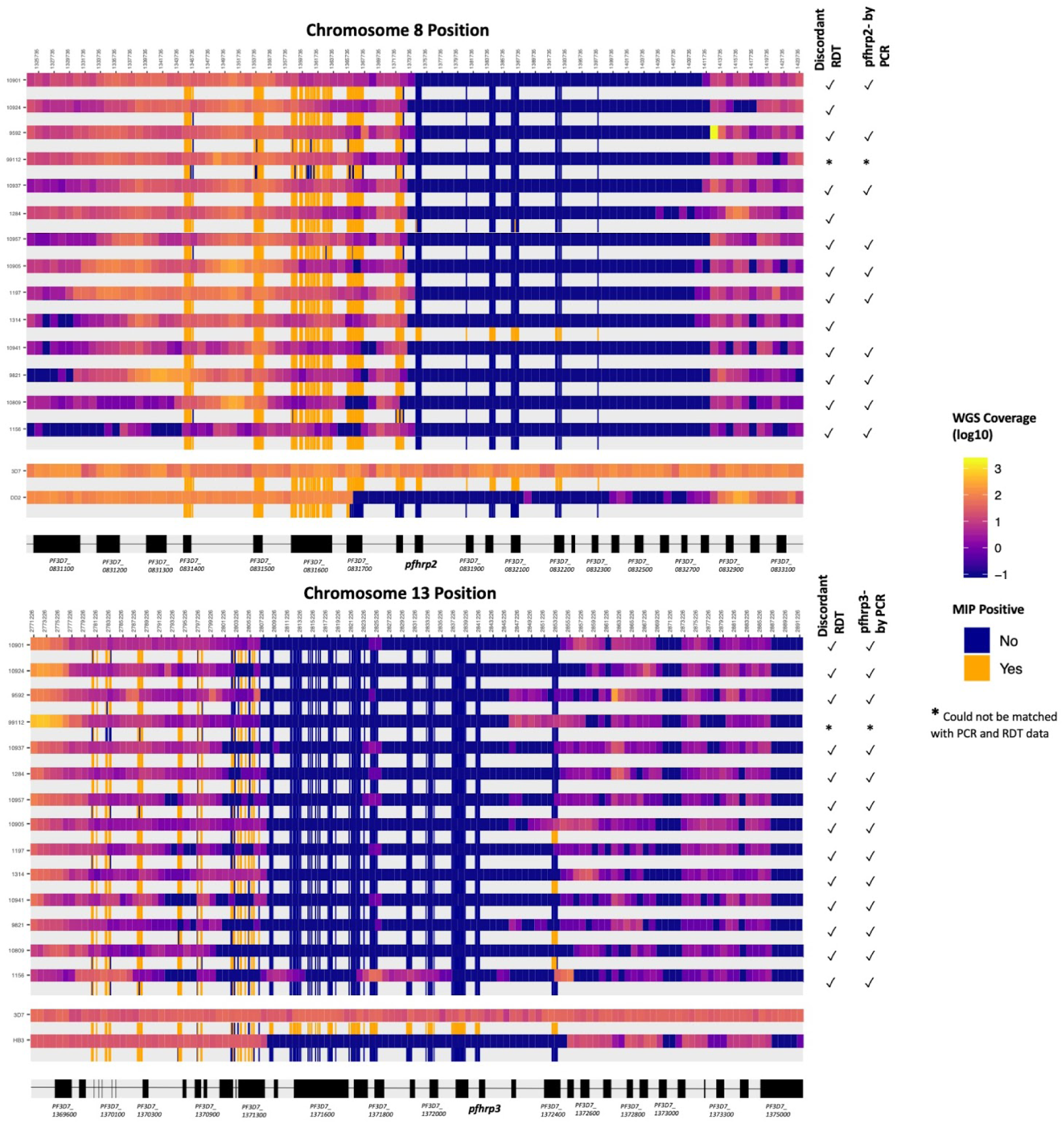
Comparison of MIP and WGS pfhrp2/3 deletion calls and breakpoint regions. Among the 14 clinical samples subjected to both methods, each sample is represented by two adjacent rows representing WGS (top) and MIP (bottom) coverage results. WGS coverage is displayed as the log_10_ median number of aligned reads per 1kb window. MIP results are colored by whether each probe captured its target, with intervening regions not targeted in the MIP panel uncolored. Sample numbers (lab_ID) are provided at left. The locations of *pfhrp2, pfhrp3*, and flanking genes are shown in black with non-genic regions in gray.

### Pfhrp2/3 deletion prevalence estimates

Incorporating RDT and PCR results, we estimated that 9.7% (95% CI 8.5-11.1) of all *P. falciparum* infections across all study sites would have false-negative HRP2-based RDT results due to *pfhrp2* deletions. Regional prevalence of false-negative RDTs due to *pfhrp2-*deleted parasites varied, with the highest estimates in Tigray (14.9%; 12.5-17.7), followed by Amhara (11.5%; 9.8-13.4) and Gambella (1.1%; 0.6-2.0). Our prevalence estimates only include samples with both the discordant RDT profile and a *pfhrp2-*negative call by PCR. Parasites with a deletion of *pfhrp2* but intact *pfhrp3* and sufficient cross-reactive HRP3 to trigger a positive HRP2 band on either RDT are not included in these estimates. Thus, the estimated prevalence of false-negative RDT results caused by *pfhrp2* deletions likely underestimates the true prevalence of *pfhrp2-*deleted parasites in this study.

### Pfhrp2/3 deletion characterization using MIP sequencing

To enable mapping of *pfhrp2/3* deletion regions and population genetic analyses in large-scale epidemiological studies, we developed a targeted panel of 241 MIPs for highly multiplexed deep sequencing of *pfhrp2, pfhrp3*, and flanking genes on chromosomes 8 and 13. A tiled design strategy was employed that involved multiple, overlapping probes spanning each gene target. We detected 244 of 273 targets with sufficient mapping quality and depth across multiple segments of both *pfhrp2* and *pfhrp3* and their flanking regions, spanning positions 1,344,451 to 1,397,773 and 2,780,863 to 2,853,533 of chromosomes 8 and 13, respectively. Fourteen total probes were used to target different segments of *pfhrp2* (n = 8 probes) and *pfhrp3* (n = 6). Among *P. falciparum* PCR-positive samples collected from 926 subjects and subjected to MIP capture and sequencing, 375 (40.5%) had sufficient depth of coverage to make high-confidence calls. In total, 43,541,045 reads were devoted to this sample set, or roughly half of a single NextSeq 550 mid-output flow cell. The median parasite density for samples successfully called using MIP sequencing was 5,077 p/µL (SD: 1.6 x 10^4^), compared to 264 p/µL (SD: 5.9 x 10^3^) for samples with failed MIP calls. Analysis of variant-called MIP sequences confirmed mixed infections with complexity of infection ≥2 in only 45 (12%) subjects; the majority (n=330, 88%) were infected by a single *P. falciparum* strain.

Among 367 (97.9%) MIP-called samples with matching PCR data, 85 (23.2%) were *pfhrp2-/3-* by PCR. MIP sequencing results indicated that 126/367 (34.3%; 95% CI 30-39) were *pfhrp2-*, 264/367 (71.9%; 67-76) *pfhrp3-*, and 116/367 (31.6%; 27-37) *pfhrp2-/3-* by MIP sequencing (**Figure 2, Table 2**). Receiver-operator curve analysis indicated the optimal parasite density threshold above which samples had sufficient coverage for MIP calling was approximately 925 p/µL, although this threshold is project-specific and is expected to improve with additional sequencing effort (**Supplementary Figure 4**).

**Figure 4.**
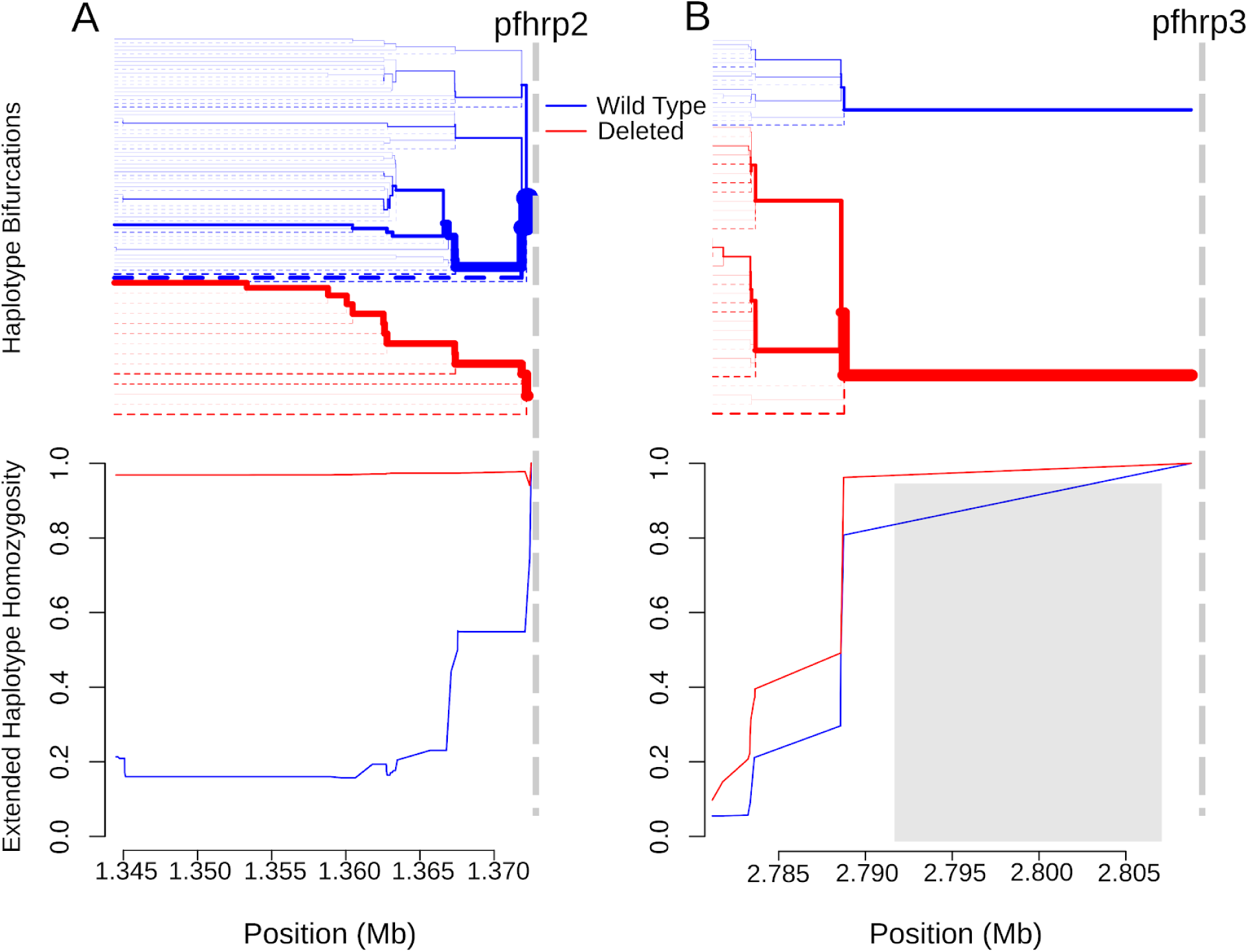
Extended haplotype homozygosity (bottom) and the bifurcation diagrams showing haplotype branching (top) centromeric to the pfhrp2 (A) and pfhrp3 (B) deletions based on MIP data. Vertical dashed lines indicate the centromeric end of deletions. No variant calls were made within the 15.5 kb region on chromosome 13 which is duplicated on chromosome 11, demarcated by the gray box (B). Abbreviations: Mb, mega-base.

### Comparison of pfhrp2/3 MIP sequencing to PCR, HRP2 immunoassay, and whole-genome sequencing results

Of samples called *pfhrp2*-by MIP sequencing, 82.0% were *pfhrp2-* by PCR and 73.3% had the discordant RDT profile. Similarly, of samples called *pfhrp3-* by MIP sequencing, 76.7% were *pfhrp3-* by PCR. While differences between genotyping results were apparent and expected due to differences in targets and methodologies (PCR is better suited for samples with low parasite density than MIP sequencing), there was overall strong concordance between RDT, PCR, and MIP results **(Table 2)**. Comparison of results from MIP and PCR *pfhrp2/3* deletion genotyping revealed excellent agreement between the two approaches for *pfhrp2* (Cohen’s kappa: 0.82) and good agreement for *pfhrp3* (Cohen’s kappa: 0.63).

Comparison to the bead-based HRP2 immunoassay results provided additional confidence in the validity of our *pfhrp2/3* deletion calls using MIP sequencing. 175 MIP-called samples also had bead-based antigen detection results available. Despite fundamental differences in the targets of these two approaches (*pfhrp2* gene versus HRP2 antigen, which can linger after clearance of infection),^32^ observed agreement between the two methods was consistent with expectation. Of those samples *pfhrp2*+ by MIP sequencing, 94.1% were HRP2+ by bead-based antigen immunoassay, whereas 79.5% of those *pfhrp2*-by MIPs were also HRP2-by the immunoassay.

We used whole-genome sequencing (WGS) to evaluate *pfhrp2/3* MIP sequencing results and breakpoint regions. Among 14 samples subjected to both WGS and MIP sequencing, median WGS depth of coverage was 20 reads/locus (range 4-38). While the distribution of aligned reads was uneven in the regions flanking *pfhrp2* and *pfhrp3*, visual inspection of WGS coverage supported 13 (93%) *pfhrp2* and 14 (100%) *pfhrp3* deletion calls made using MIP sequencing data (**Figure 3**). For the single discordant *pfhrp2* deletion call (lab ID: 1314), *pfhrp2* PCR results were consistent with the MIP sequencing result. Precise mapping of breakpoint regions using WGS was not possible due to regions of very high coverage (“jackpotting”) resulting from selective whole-genome amplification and low coverage due to ambiguous read mapping to repetitive and paralogous loci. However, breakpoint regions identified using MIPs were consistent with WGS coverage centromeric to *pfhrp2* and *pfhrp3* on chromosomes 8 and 13, respectively, with the exception of calls in chromosome 13’s multi-copy 28S rRNA gene. Discordance in these calls was expected due to ambiguous mapping of short-read sequences to a multi-copy gene. MIP results from well-characterized lab strains 3D7 (*pfhrp2+/3+*), DD2 (*pfhrp2-/3+*), and HB3 (*pfhrp2+/3-*) were consistent with whole-genome alignments of published short-read data. Telomeric deletion breakpoint assessment was limited by a small number of successful MIP targets telomeric to both genes. However, the concordance in *pfhrp2/3* deletion calls and centromeric deletion breakpoint regions by MIP and WGS techniques confirmed the utility of MIPs for identifying *pfhrp2/3* deletions and determining their extent and breakpoint regions.

### Pfhrp2/3 deletion breakpoint profiling

Compared to PCR, bead-based immunoassay, or RDT diagnosis, MIP sequencing was unique in its ability to reveal distinct subtelomeric structural profiles along chromosomes 8 and 13 into which samples could be categorized: three for *pfhrp2*+ samples (chr8-P1, chr8-P2, chr8-P3), one for *pfhrp*2-(chr8-P4), one for *pfhrp3*+ (chr13-P1), and three for *pfhrp3*-(chr13-P2, chr13-P3, chr13-P4) (**Figure 2**). All *pfhrp2-* samples had the same subtelomeric structural profile (chr8-P4), although two other subtelomeric deletions were identified on chromosome 8 that did not involve *pfhrp2* (chr8-P2, chr8-P3). These deletions involve members of the *rifin* and *stevor* gene families, as well as genes of unknown function.

The structural profile of most samples identified as *pfhrp3-* (chr13-P3 and chr13-P4) differed in the presence or absence of a segment of chromosome 13 directly telomeric to *pfhrp3* (position 2,852,540 - 2,853,533) encoding a member of the acyl-coA synthetase family (PF3D7_1372400). All chr13-P3 and chr13-P4 deletions resulted in loss of genes with roles in red blood cell invasion (PF3D7_1371700, serine/threonine kinase and member of the FIKK family; PF3D7_1371600, erythrocyte binding-like protein 1 [*EBL-1*]),^33,34^ while they were present in all chr13-P1 (*pfhrp3-*intact) parasites. The chr13-P2 deletion profile was observed in only one sample from Amhara’s Metema district. We did not observe an association between subtelomeric structural profile and the number of symptoms experienced by subjects (**Supplementary Figure 5, Supplementary Results**) or geographic region (**Supplementary Table 4**).

Analysis of 25 genomes from *P. falciparum* samples collected in Ethiopia in 2013 and 2015 and available in the MalariaGEN database (**Supplementary Figures 6-7**) uncovered chromosome 13 subtelomeric structural profiles similar to those identified by MIP sequencing: 9 samples with coverage consistent with chr13-P3 (*pfhrp3-*deleted), 2 samples with chr13-P4 (*pfhrp3-*deleted), and 14 samples with chr13-P1 (*pfhrp3-*intact).^19^

### Genetic signatures of evolutionary selection

Extended haplotype homozygosity (EHH) statistics revealed signatures of recent positive selection in the flanking region centromeric to *pfhrp2* deletions on chromosome 8 but not in flanking regions around *pfhrp3* deletions on chromosome 13^35^. 91 and 17 biallelic SNPs within the 28kb and 27kb regions centromeric to *pfhrp2* and *pfhrp3* deletions, respectively, were used to calculate EHH statistics. 327 samples with *pfhrp2* deletion calls using MIP sequencing and sufficient variant data were included in the EHH analysis, including 212 *pfhrp2-*intact and 115 *pfhrp2*-deleted haplotypes. EHH remained very high for parasites with the *pfhrp2* deletion (0.968) along the entire 28kb analyzed, whereas homozygosity around the *pfhrp2-*intact (wild-type) allele quickly broke down (**Figure 4A**). A similar pattern was observed when deletion profiles were analyzed separately; chr8-P4 EHH remained high and chr8-P1-P3 EHH quickly broke down (**Supplementary Figure 8**).

We further confirmed high EHH around the *pfhrp2* deletion allele using WGS data. Comparing 23 whole-genome sequenced samples from this study and the 25 published MalariaGEN samples described above, we were able to extend our analysis and confirm an EHH length of >143kb centromeric to the deletion **(Supplementary Figure 9A)**. These findings suggest a recent selective sweep, indicative of strong evolutionary pressure favoring *pfhrp2-*deleted *P. falciparum* parasites.

A different pattern was observed in the regions flanking *pfhrp3* (**Figure 4B**). 162 samples with *pfhrp3* deletion calls using MIP sequencing and sufficient variant data were included in the EHH analysis, including 37 *pfhrp3-*intact and 125 *pfhrp3*-deleted haplotypes with three distinct subtelomeric structural profiles. No variant calls were made in the 15.5 kb region immediately centromeric to *pfhrp3* to avoid ambiguity in read mapping to the duplicated DNA segment containing multicopy genes including 5.8S, 28S rRNA. EHH quickly decreased below 0.5 for *pfhrp3* deletion alleles as well as the *pfhrp3-*intact allele within 1 kb of available SNPs. When deletion profiles were analysed as separate alleles, the EHH pattern was similarly low for chr13-P1, P3 and P4 (**Supplementary Figure 10**). Comparison of EHH around the *pfhrp3*-intact and P3-like *pfhrp3* deletions using WGS data from the 25 MalariaGEN samples confirmed our finding that the EHH quickly decreased for both *pfhrp3-*intact and *pfhrp3-*deletion alleles (**Supplementary Figure 9B**). Taken together, these findings suggest that each *pfhrp3* deletion profile arose multiple times independently, and/or they have been present in the parasite population for sufficient time for homozygosity due to genetic hitchhiking to be degraded by recombination with different haplotypes.

## DISCUSSION

Using the largest prospective study of *pfhrp2/3-*deleted *P. falciparum* performed to-date and complementary molecular, immunological, and novel sequencing assays, we provide clear evidence that *pfhrp2/3-*deleted parasites are circulating in multiple sites along Ethiopia’s borders with Sudan and Eritrea. Analysis of flanking haplotypes suggests that the *pfhrp2* deletion mutation emerged and recently expanded from a single origin, while *pfhrp3* deletion mutations have existed for a longer time span and likely have multiple origins. As expected, we did not observe perfect concordance between RDT results, PCR, a bead-based immunoassay, WGS, and MIP sequencing results. However, the preponderance of evidence from these diverse platforms provides robust confirmation of deletions and supports the use of World Health Organization (WHO) protocols for rapid *pfhrp2/3* deletion surveillance.^29^ The prevalence of false-negative HRP2-based RDT results due to *pfhrp2* deletions is estimated at 9.7% overall and up to 11.5% and 14.9% in the Amhara and Tigray regions, respectively. These estimates exceed WHO minimum criteria (>5%) for a change in national diagnostic testing strategy. *Pfhrp2/3-*deleted parasites threaten recent progress made by Ethiopia’s malaria control and elimination program, and raise concerns about ongoing use of and exclusive reliance on HRP2-based RDTs in the region for diagnosis of falciparum malaria.

Eritrea’s alarming reports of false-negative RDTs due to *pfhrp2/3-*deleted parasites prompted an immediate change in national diagnostic testing policy in 2016^15,36^. Recent evidence from Sudan, Djbouti, and Somalia suggests that the Horn of Africa may already be heavily affected by *pfhrp2/3-*deleted parasites^37,38^, though results from ongoing surveillance efforts are not yet publicly available. Within affected regions in Ethiopia, we observed spatial heterogeneity in *P. falciparum* RDT profiles by district, with prevalence of the discordant HRP2-, PfLDH+ RDT profile ranging from 0.9 to 30% (**Supplementary Table 2**). While finer scale spatial analyses were not possible due to our health facility sampling approach, this finding is consistent with prior studies showing variation within countries and by region^25^. Differences in transmission intensity, treatment-seeking behavior, diagnostic testing capacity, and seasonality may account for some of the spatial variation in *pfhrp2/3* deletion prevalence estimates^17,39–41^. Although the factors driving emergence of these parasites in some regions but not others remain poorly understood, our study suggests that *pfhrp2-*deleted parasites may have spread widely within Ethiopia from a single origin. This finding is consistent with early microsatellite analysis of *pfhrp2/3-*deleted strains in Eritrea, in which 30 of 31 (96.8%) *pfhrp2-*deleted strains fell into a single genetically related cluster,^15^ and raises concern about clonal expansion of *pfhrp2-*deleted strains in the Horn of Africa.

Using a multi-faceted approach, we validate the use of MIP sequencing for high-throughput *pfhrp2/3* deletion genotyping, deletion profiling, and population genetic analysis. Comparison of MIP sequencing to other approaches demonstrated that it can be used for cost-effective (approximately $10-15 per sample) and scalable deletion genotyping in samples with parasite densities of approximately 1,000 parasites/µL. While this threshold can likely be improved by additional sequencing of samples with inadequate sequencing depth-of-coverage, in this case, the equivalent of half a NextSeq 550 flow cell enabled visualization of deletion breakpoint regions and variant calling in *P. falciparum’*s subtelomeres in a large portion of samples, without the need for costly enrichment and WGS.

Based on analysis of MIP sequencing and available WGS data, we posit one potential model by which *pfhrp2/3-*deleted parasite populations may have evolved in the Horn of Africa. Findings from this study suggest that parasite populations with *pfhrp3* deletions expanded in the more distant past and potentially arose multiple times independently, based on low EHH surrounding *pfhrp3*, multiple deletion profile patterns, the high overall frequency of *pfhrp3-*deleted parasites, and their presence in older samples from 2013 in the MalariaGEN study. In this milieu, recent strong selection favoring parasites with deletions of *pfhrp2* likely occurred due to “test-track-treat” policies that rely upon HRP2-based RDTs and allow parasites with deletions of both genes, or in some cases one of the two genes, to escape treatment. Implicit in this model is the assumption that forces apart from RDT-derived pressure are also driving the evolution of *pfhrp2/3* deletions. First, test-track-treat policies do not explain the prevalence and genetic evidence of well-established *pfhrp3* deletions in Ethiopia, as malaria due to *pfhrp2+/3-* parasites should be detectable by HRP2-based RDTs^42^. Second, *pfhrp2/3-*deleted parasites are highly prevalent in South America,^25^ where RDT-based treatment decisions have never been common. Third, the prevalence of *pfhrp2/3-*deleted parasites appears to have remained stable in Eritrea despite removal of HRP2-based RDTs two years ago.^38^

What other advantages might *pfhrp2/3-*deleted parasites have over those with intact genes? Our limited understanding of the biology of these deletions makes this question hard to answer. Several lines of inquiry may be relevant: 1) They may be better adapted to low transmission intensity settings than other strains. *Pfhrp2/3-*deleted parasites appear to be more common in regions with lower transmission and, presumably, lower complexities of infection.^40^ This trend might simply be an artifact of the assays used to detect them -i.e., neither PCR, antigen immunoassays, nor common sequencing methodologies are well suited to detect a *pfhrp2/3-*deleted strain when *pfhrp2/3-*intact strains have co-infected a human host. The high frequency of monoclonal samples in our MIP sequence analysis provides support for this hypothesis. However, it is also possible that the *pfhrp2/3* genes are an asset when within-host competition is common but a liability when it is rare. This makes sense if parasites in low-transmission settings gain an advantage through improved gametocytogenesis and transmission, for example. 2) Loss of *pfhrp2/3* or flanking genes may alter parasite virulence. Evidence is accumulating that HRP2 plays a role in cerebral malaria and endothelial inflammation during severe malaria^43,44^. People infected by *pfhrp2/3-*deleted parasites may have less severe disease and therefore be less likely to seek treatment, increasing the likelihood of onward transmission. However, we cannot exclude the possibility that *pfhrp2/3* are lost as a consequence of selection on other genes. For example, the flanking gene *EBL-1* is almost uniformly lost in *pfhrp3-*deleted parasites in this cohort and appears to play a role during invasion of red blood cells^34,45^. Similarly, members of the *rifin* and *stevor* gene families with potential roles in parasite virulence were lost in the subtelomeric deletions observed in this study^46,47^. We did not observe evidence of an association between virulence and subtelomeric deletions in our cohort, but limited clinical data prevents us from assessing the hypothesis rigorously. 3) Loss of *pfhrp2/3* or flanking genes may improve transmissibility to or from mosquitoes. To our knowledge, this phenomenon has not been studied. These and other hypotheses require experimental and improved epidemiological analyses. Regardless of the evolutionary forces at play, our findings strongly suggest that the evolution of *pfhrp2/3-*deleted parasites in Ethiopia was a multi-step process that involved earlier expansion of *pfhrp3-* than *pfhrp2-*deleted parasite populations.

This study has several limitations. First, the study design prioritized evaluation of samples with discordant RDT results (HRP2-but Pf-LDH+) for rapid assessment of false-negative RDTs due to *pfhrp2/3* deletions in the context of clinical treatment. This feature of the WHO protocol is intentional as it captures clinically significant *pfhrp2/3* deletions and enables real-time, efficient signaling to malaria control programs of a potential problem, but it also introduces selection bias that requires careful consideration when estimating the true prevalence of *pfhrp2/3-*deleted parasites. We overcame this limitation by using a conservative approach that incorporated both RDT and PCR data to estimate false-negative RDT results due to *pfhrp2* deletions. This metric is relevant to control programs, but does not capture asymptomatic or low-parasite-density infections by *pfhrp2/3-*deleted parasites. Second, only a subset of samples underwent advanced analysis, and clinical data was not available for all subjects. This was not unexpected for a pragmatic field study of this size. We do not believe that it introduced sufficient bias into our prevalence estimates or population genetic analysis to change our conclusions. Third, we cannot comment on changes in selection pressure over time because the study was cross-sectional. Fourth, we only sampled three regions of Ethiopia, which is a diverse and populous country. In response, the Federal Ministry of Health is now conducting a country-wide survey that will enable comparison of *pfhrp2/3* deletions over time in select sites.

## CONCLUSION

Leveraging a large prospective study, established molecular and antigen detection methods, and a novel targeted sequencing approach, we demonstrate that *pfhrp2/3-*deleted *P. falciparum* is a common cause of false-negative RDT results among subjects presenting with symptomatic malaria in three regions of Ethiopia. The genomic tools employed in this study reveal complex origins of these parasites. Recent, strong selective pressures favoring *pfhrp2-*deleted parasites appear to have occurred on a background of pre-existing *pfhrp3* deletions. Existing malaria control programs in the region are threatened by expansion of these parasite strains, and surveillance is urgently needed to inform decisions about when alternative malaria diagnostics should be deployed.

## METHODS

### Study Design and Data Collection

We performed a cross-sectional, multi-site study in eleven districts along Ethiopia’s borders with Eritrea, Sudan, and South Sudan, located within three of its nine administrative regions. On average, ten health facilities were selected from each district, including four districts of Amhara Region (northwest Ethiopia), six districts of Tigray Region (north Ethiopia), and one district of Gambella Region (southwest Ethiopia) during the 2017-2018 peak malaria transmission season (September-December) (see **Figure 1**). Per WHO protocol^29^, each facility passively enrolled participants presenting with symptoms of malaria (fever, headache, joint pain, feeling cold, nausea, and/or poor appetite), with sample size proportionally allocated to each facility based on the previous year’s malaria case load. All participants provided informed consent, participated in interview questionnaires, and underwent blood collection for RDT testing using two types of RDTs. Ethical approval was obtained from the Ethiopia Public Health Institute (EPHI) Institutional Review Board (IRB; protocol EPHI-IRB-033-2017) and WHO Research Ethics Review Committee (protocol: ERC.0003174 001). Processing of de-identified samples and data at UNC was determined to constitute non-human subjects research by the UNC IRB (study 17-0155). The study was determined to be non-research by the Centers for Disease Control and Prevention Human Subjects office (0900f3eb81bb60b9). Experiments were performed in accordance with relevant guidelines and regulations.

### Field Sample Evaluation

Study participants were evaluated using both a CareStart *Pf/Pv* (HRP2/Pv-pLDH) RDT (Access Bio, Somerset, NJ; product code RM VM-02571) and an SD Bioline Malaria Ag P.f. (HRP2/Pf-LDH) RDT (Alere, Waltham, MA; product 0code 05FK90). For the CareStart RDT, 5 µL of capillary whole blood was collected by finger prick and transferred to the RDT sample well, along with 60 µL of buffer solution. Results were read at 20 minutes. The SD Bioline RDT followed the same protocol, but with 4 drops of buffer added and results read in a 15-30 minute window. Participants testing positive by either RDT were first prescribed treatment, according to Ethiopian national guidelines.^48^

Cases with any positive HRP2 or Pf-LDH RDT band were considered positive for *P. falciparum* malaria. Cases Pf-LDH-positive but HRP2-negative on both RDTs were considered potential candidates for *pfhrp2/3* gene deletion and defined as ‘discordant.’ These participants, along with a subset of HRP2-positive and -negative controls, provided further informed consent for additional blood collection for dried blood spot (DBS) preparation. At least two DBS samples (50 µL/spot) were collected on Whatmann 903 protein saver cards (GE Healthcare, Chicago, IL) from consenting participants. DBS were stored in plastic bags with desiccant. A randomly selected subset of DBS were sent for molecular analysis to the University of North Carolina at Chapel Hill and for serological analysis to the Centers for Disease Control and Prevention (CDC, Atlanta, GA).

### DNA Extraction and PCR assays

DNA was extracted from three 6mm punches per DBS sample using Chelex-100 and saponin as previously described^49^. Quantitative PCR (qPCR) assays were first performed in duplicate for *pfldh*^*50*^. To avoid the risk of misclassification due to DNA concentrations below the limit of detection for *pfhrp2/3* PCR assays, further analysis was restricted to samples with >100 parasites/µL by qPCR (**Supplementary Figure 2**).^14^ PCR assays targeting exon 2 of *pfhrp2* and *pfhrp3* were then performed in duplicate as previously described,^7^ except that PCRs were performed as single-step, 45-cycle assays, using 10µL template and AmpliTaq Gold 360 Master Mix (Thermo Fisher Scientific, Waltham, MA) in 25 µL reaction volume. In addition to no-template and *P. falciparum* 3D7 strain (*pfhrp2+/3+*) positive controls, *pfhrp2* assays included an additional DD2 strain (*pfhrp2-/3+*) control and *pfhrp3* assays included an additional HB3 strain (*pfhrp2+/3-*) control. Finally, an additional single-copy gene, real-time PCR assay targeting *P. falciparum* beta-tubulin was performed to confirm that sufficient parasite DNA remained in samples with a negative *pfhrp2/3* PCR result.^14^ *Pfhrp2/3* genotyping calls were made in samples with *pfldh* qPCR parasitemia >100 parasites/µL to avoid misclassification in the setting of amplification failure due to low target DNA concentration. A *pfhrp2* or *pfhrp3* positive call required ≥1 replicate with distinct band(s) with expected fragment length. A negative call required both *pfhrp2* or *pfhrp3* replicates to be negative. Detailed reaction conditions for all PCR assays are described in the **Supplementary File**.

### Serological assays

The presence of HRP2, pan-LDH, and aldolase antigenemia was assessed in a subset of DBS samples (single 6mm punch) using a multiplex bead-based immunoassay as previously described^13^. Within this multiplex assay, capture and detection antibodies against the HRP2 antigen would also recognize similar epitopes on the HRP3 antigen, so unique signals for these two antigens cannot be obtained.

### Prevalence estimates

We estimated the prevalence of *P. falciparum* infections expected to have false-negative HRP2-based RDT results due to *pfhrp2* deletions as follows. First, we calculated the proportion of all RDT-positive *P. falciparum* cases (HRP2+ or PfLDH+ on any RDT) with the discordant RDT profile (HRP2- on both RDTs, but PfLDH+), overall and by region. Second, we calculated the observed concordance between the discordant RDT profile and a *pfhrp2-*negative PCR call, overall. Prevalence estimates and 95% CIs were then back-transformed overall and by region using the ci.impt function within the *asbio* R package, which generates CIs for the product of two proportions using delta derivation. This allowed us to estimate with confidence the proportion of *P. falciparum* infections with both *pfhrp2* deletions and false-negative HRP2-based RDT results, overall and by region. As a sensitivity analysis, we also estimated the proportion of those with a discordant RDT and a *pfhrp2-*negative PCR call (directly multiplying the true proportion of Pf-positive individuals with a discordant RDT profile, overall and by region, by 0.727, or the overall proportion of discordant RDT samples that had a *pfhrp2-* PCR result). 95% CIs were then generated using bootstrapping (1000 iterations). The prevalence estimates and CIs generated by the two approaches were similar **(Supplementary Table 3)**.

### Pfhrp2/3 molecular inversion probe (MIP) development

*Pfhrp2, pfhrp3*, and the flanking regions within a 100kb window surrounding each gene were targeted for MIP designs using *MIPTools*^*51*^. A tiled design strategy was employed that involved multiple, overlapping probes spanning each gene target. 22 genes flanking *pfhrp2* and 31 genes flanking *pfhrp3* were used in the design, of which 11 and 19 were successful on the first design try, respectively. A second attempt was not made for designs for the flanking genes. A total of 241 probes; 9 for *pfhrp2*, 9 for *pfhrp3* and 223 probes for the flanking genes were designed. MIPs were designed using the 3D7 (v3) reference genome avoiding hybridization arms in variant regions when possible. 80 alternative probes accommodating potential variants in the highly variable *pfhrp2* and *pfhrp3* genes were also created. A 15.5 kb segment centromeric to *pfhrp3* on chromosome 13 between positions 2792000-2807500 is duplicated on chromosome 11 between positions 1918007-1933488, with 99.4% sequence identity. Therefore, the target genes falling into this region were multicopy genes and their probes were designed to bind to both loci on the genome (see **Supplementary Table 5** for the design overview including all genes targeted, MIPs designed and genomic coordinates). Probes were ordered from Integrated DNA Technologies (Coralville, Indiana, USA) as 200 pmol ultramer oligos. Probe sequences are provided in the **Supplementary Table 6**.

### MIP capture and deep sequencing of clinical samples

All DNA samples extracted by UNC underwent MIP capture using the capture and amplification methods exactly as described by Verity *et al*.^52^, with the exception of oligonucleotides (the *pfhrp2/3* MIP oligonucleotide panel described above was used) and controls (we selected a different set of controls that are informative for *pfhrp2/3* deletion characterization). All MIP captures included multiple controls: 3D7 (*pfhrp2+/3+*), DD2 (*pfhrp2-/3+*), HB3 (*pfhrp2+/3-*) laboratory strains; as well as LC and HC mixes (1% HB3, 10% DD2, 89% 3D7) at 250 and 1000 parasites/µl densities, respectively. Samples were sequenced on the Illumina NextSeq 550 instrument using 150bp paired-end sequencing and dual indexing.

### Subtelomeric profiling and variant calling with MIP data

Read mapping and variant calling were carried out using *MIPTools* (v0.19.12.13)^51^. *MIPTools* uses the *MIPWrangler* algorithm (v1.2.0)^53^ to create high quality consensus sequences from sequence read data utilizing unique molecular indexes (UMIs) of MIPs, maps those sequences to the reference genome using *bwa* (v0.7.17) and remove off target sequences as described previously^31,52^. Deletion calls were limited to the samples that had high coverage to avoid false positives. Considering the high frequency of large deletions present in the sample set, the coverage threshold was based on a subset of probes that were present on > 60% of the samples, none of which overlapped with the chromosome 8 or 13 deletions. Samples with a median coverage of < 5 UMIs for this subset of probes were excluded from analysis.

Structural profiling was performed using the UMI count table (**Supplementary Table 7**). The count table was converted to a presence/absence table such that if a probe had > 1 UMI for a given sample, it was accepted as present (i.e., not deleted). Samples were clustered into subtelomeric structural profile groups based on this table using the hierarchical clustering algorithm AgglomerativeClustering of the Python module Scikit-learn (v0.20)^54^ using only the regions involved in the deletion events of the corresponding chromosome (position > 1372615 for chromosome 8 and position > 2806319 for chromosome 13). Samples were grouped into their final subtelomeric structural profile based on visual inspection of the resulting clusters.

Initial variant calls were made using *freebayes* (v1.3.1) via *MIPTools* with the following options: --pooled-continuous --min-base-quality 1 --min-alternate-fraction 0.01 --min-alternate-count 2 --haplotype-length −1 --min-alternate-total 10 --use-best-n-alleles 70 --genotype-qualities. Variants were processed using *MIPTools* to filter for: variant quality > 1, genotype quality > 1, average alternate allele quality > 15, minimum depth > 2 UMIs; and make final genotype calls based on the major allele (within sample allele frequency > 0.5). In addition, the following variants were removed from the final call set: those that were observed as a major allele in less than two samples (singletons), not supported by more than two UMIs in at least three samples, present on multicopy genes, and indels. Variant calls were further filtered for missingness to avoid imputation in EHH calculations: samples missing calls for >50% of the variants were removed, variants missing calls in >50% of the samples were removed. Variants calls were converted to .map and .hap files (**Supplementary Table 8**) for use with the *rehh* package in R.

### Assessment of MIP calls using whole-genome sequencing

We performed WGS on a subset of samples to assess the accuracy of MIP *pfhrp2/3* deletion calls. DNA extracted from samples with discordant RDT results were selected for *P. falciparum* selective whole-genome amplification (sWGA) and whole-genome sequencing as described previously^30^. In brief, DNA was first subjected to two separate sWGA reactions using the Probe_10 primer set described by Oyola et al.^55^ and the JP9 primer set.^30^ sWGA products were then pooled in equal volumes and acoustically sheared using a Covaris E220 instrument prior to sequencing library preparation using Kappa Hyper library preps. Indexed libraries were then pooled and sequenced on an Illumina HiSeq 4000 instrument using 150bp, paired-end sequencing. Sequencing reads were deposited into NCBI’s Sequence Read Archive (accession numbers pending).

### Published whole genome sequencing data retrieval

Fastq files from 25 Ethiopian samples included in the MalariaGEN genome variation project^19^ and 3 laboratory strains (3D7, HB3 and DD2) from MalariaGEN genetic crosses project^56^ were downloaded from the European Nucleotide Archive using fasterq-dump (v2.10.8) and sample accession numbers (**Supplementary Table 9**).

### WGS data analysis

All fastq files were processed as follows. Adapter and quality trimming was performed using *Trimmomatic* (v.0.39) with the recommended options (seed mismatches:2, palindrome clip threshold:30, simple clip threshold:10, minAdapterLength:2, keepBothReads LEADING:3 TRAILING:3 MINLEN:36). Trimmed fastq files were mapped to 3D7 reference genome (v3.0) concatenated to human genome (hg38) to avoid incorrect mapping of reads originating from host DNA using *bowtie2* (v2.3.0) with the ‘--very-sensitive’ option. Reads mapping to the parasite chromosomes were selected and optical duplicates were removed using the *sambamba* (v0.7.1) view and markdup commands, respectively. Read coverage was calculated using *samtools* (v1.9) depth command with options ‘-a -Q1 -d0’, filtering reads with mapping quality of zero. Variants were called only for the regions of interest using *freebayes* (v1.3.1) with the following options: ‘--use-best-n-alleles 70 --pooled-continuous --min-alternate-fraction 0.01 --min-alternate-count 2 --min-alternate-total 10 --genotype-qualities --haplotype-length −1 --min-mapping-quality 15 -r region’. Regions of interest were from 300 kb centromeric to the deletions to chromosome ends (positions 1074000-1472805 and 2505000-2925236 for chromosomes 8 and 13, respectively).

Variants were filtered for: variant quality > 20, genotype quality > 15, average alternate allele quality > 15, minimum depth > 4 reads. In addition, the following variants were removed from the final call set: those that were never observed as a major allele in any sample, not supported by more than 10 reads in at least one sample, and indels. Final genotype calls were based on the major allele (within sample allele frequency > 0.5). Variant calls were further filtered for missingness to avoid imputation in EHH calculations: samples missing calls for >95% of the variants were removed, variants missing calls in >10% of the samples were removed. Telomeric profiling of the published genomes was carried out by visual inspection of depth-of-coverage plots (**Supplementary Figures 8** and **9**). Summary statistics were generated (**Supplementary Table 10**) using the python pandas module (v0.23).

### Statistical and population genetic analysis

Data collected during the participant’s study visit (clinical data and RDT results) was linked to laboratory results via the barcode number transcribed on DBS sent to the UNC and CDC laboratories. Samples in the dataset with missing or duplicate barcodes were arbitrated using original paper questionnaires by the EPHI data center. Ultimately, an analysis dataset that included both PCR and field data was created including all samples we could confidently merge by both barcode number and region label.

Statistical analysis was performed using R (version 3.6.0, R Core Team, Vienna, Austria, 2019; www.R-project.org). ArcGIS (Desktop Version 10.5, ESRI, Redlands, CA, 2016) was utilized for mapping, with additional annotation performed using PowerPoint (version 16.31, Microsoft, Redmond, WA, 2019).

Extended haplotype homozygosity (EHH) statistics were calculated to evaluate the regions flanking the *pfhrp2* and *pfhrp3* genes for signatures of recent positive selection^35^ using the *rehh* package (version 3.1.2)^57^. EHH statistics were calculated using the data2haplohh and calc_ehh functions, haplotype furcations were calculated using calc_furcation, and plots were generated using the package’s plot function and annotated using Inkscape (version 0.92).

Complexity of infection (COI) for each sample was calculated using *McCOILR* (v1.3.0, https://github.com/OJWatson/McCOILR), an *Rcpp* wrapper for *THE REAL McCOIL*^*58*^ with the options maxCOI=25, totalrun=2000, burnin=500, M0=15, err_method=3. The same variant set used in the EHH analysis was used for the COI calculations, except that variants whose within-sample allele frequency were between 0.05 and 0.95 were called heterozygote for COI analysis.

## Supporting information

Supplementary File - PCR conditions

## Data Availability

Genomic sequencing data will be available through the Sequence Read Archive (BioSample accession numbers pending). De-identified datasets generated during the current study will be available as supplementary files. Code used during data analysis will be made available on GitHub.

## ACKNOWLEDGMENTS

The authors thank the research teams for conducting field work and the subjects for participating in the study. They would also like to thank Steven R. Meshnick posthumously for contributions to the laboratory analyses and data analysis. The following reagents were obtained through BEI Resources, NIAID, NIH: Genomic DNA from *P. falciparum* strain 3D7, MRA-102G, contributed by Daniel J. Carucci; *P. falciparum* strain HB3, MRA-155G, contributed by Thomas E. Wellems; *P. falciparum* strain Dd2, MRA-150G, contributed by David Walliker. The authors also would like to acknowledge MSF Holland for supporting the field study in Gambella region.

## Author contributions

SMF, JAC, and JBP conceived the study. SMF, HM, BGB, HM, HS, BP, EA supervised and/or conducted field work. OA, CH, MD, ER performed laboratory assays and experiments. ENR, OA, CK, JJJ, JAB, ER, JBP analyzed laboratory data. ENR and OA produced the tables and figures. SMF and ENR wrote the first draft with assistance from OA and JBP. All authors critically reviewed and approved the final manuscript.

## Disclaimer

The findings and conclusions in this report are those of the authors and do not necessarily represent the official position of the CDC.

## SUPPLEMENTARY MATERIAL

### Supplementary Results

#### Association between malaria symptoms, geographical location, and subtelomeric structural variants

We did not observe an association between subtelomeric deletion profile and the number of symptoms experienced by subjects (**Supplementary Figure 5**). Because the majority of subjects (96.5%, 2620/2714) who tested positive for *P. falciparum* by RDT were febrile, fever alone was not sufficient to evaluate disease severity. Therefore, as a crude metric of disease severity, we calculated the total number of symptoms (six total were assessed: fever, headache, joint pain, feeling cold, nausea, and lack of appetite). No significant differences in the total number of symptoms by deletion profile were revealed for chromosome 8 (one-way ANOVA p = 0.83) or chromosome 13 (p = 0.72). No obvious spatial patterns in subtelomeric deletion profiles were apparent at the regional level (**Supplementary Table 2**).

#### Comparison of pfhrp2/3 PCR and HRP2 bead-based assays

We observed expected agreement between the results of *pfhrp2/3* PCR assays, RDTs, and a bead-based immunoassay applied to a subset of 456 samples. 93% (95% CI 86-96) of samples *pfhrp2+/3+* by PCR tested positive for HRP2 antigen (GM 40,284 pg/mL HRP2, geoSD 7.5). In comparison, 19% (12-29) of *pfhrp2-/3-* samples were HRP2+, with a GM of 2,089 pg/mL HRP2 (geoSD 5.5). HRP2+ but *pfhrp2-* PCR results are expected in a subset of subjects because HRP2 antigenemia can persist for weeks after clearance of parasitemia^32^. 92% (95% CI 88-95) of samples HRP2+, Pf-LDH+ by RDT were HRP2+ by the antigen assay (GM 34,536 pg/mL, geoSD 6.5), compared to 24% (95% CI 18-31) of those with the discordant HRP2-, Pf-LDH+ RDT profile of interest (GM 2,455 pg/mL, geoSD: 7.2) (**Table 2**).

#### Subtelomeric profiling and variant calling using MIPs

241 MIPs mapped to 273 targeted loci on the reference genome, including 32 extra loci accounting for the multicopy genes on chromosome 11. Probes failing to amplify in >90% of the samples were removed from the analysis, leaving 244 loci. 841 of 1014 samples and controls had sequence data after read mapping. 20 of 841 belonged to control strains (positive controls). None of the 20 negative controls had any sequence mapping to the reference genome. Deletion calls were only made in samples with sufficient depth of UMI coverage (see Methods), leaving 375 high-coverage samples from the study cohort and 6 controls in the final call set.

### Supplementary Files

*Supplementary tables are compiled into a single file for ease of viewing*.

**Supplementary Table 1**. PCR results by age, sex, and parasite density.

**Supplementary Table 2**. RDT profile by district for individuals *P*. *falciparum*-positive by RDT.

**Supplementary Table 3**. Prevalence estimate sensitivity analysis.

**Supplementary Table 4**. MIP subtelomeric structural profiles by region.

**Supplementary Table 5**. *Pfhrp2/3* MIP panel design overview, including genes

targeted, MIPs designed, and genomic coordinates.

**Supplementary Table 6**. *Pfhrp2/3* MIP panel probe sequences.

**Supplementary Table 7**. Absolute (A) and normalized (B) unique molecular identifier (UMI) counts by sample and locus.

**Supplementary Table 8**. MIP variant calls, which were converted to .map and .hap files.

**Supplementary Table 9**. ENA accession numbers of previously published WGS data.

**Supplementary Table 10**. Summary statistics of WGS coverage for samples sequenced in this study.

**Supplementary File**. PCR reaction conditions.

## Supplementary Figures

**Supplementary Figure 1.**
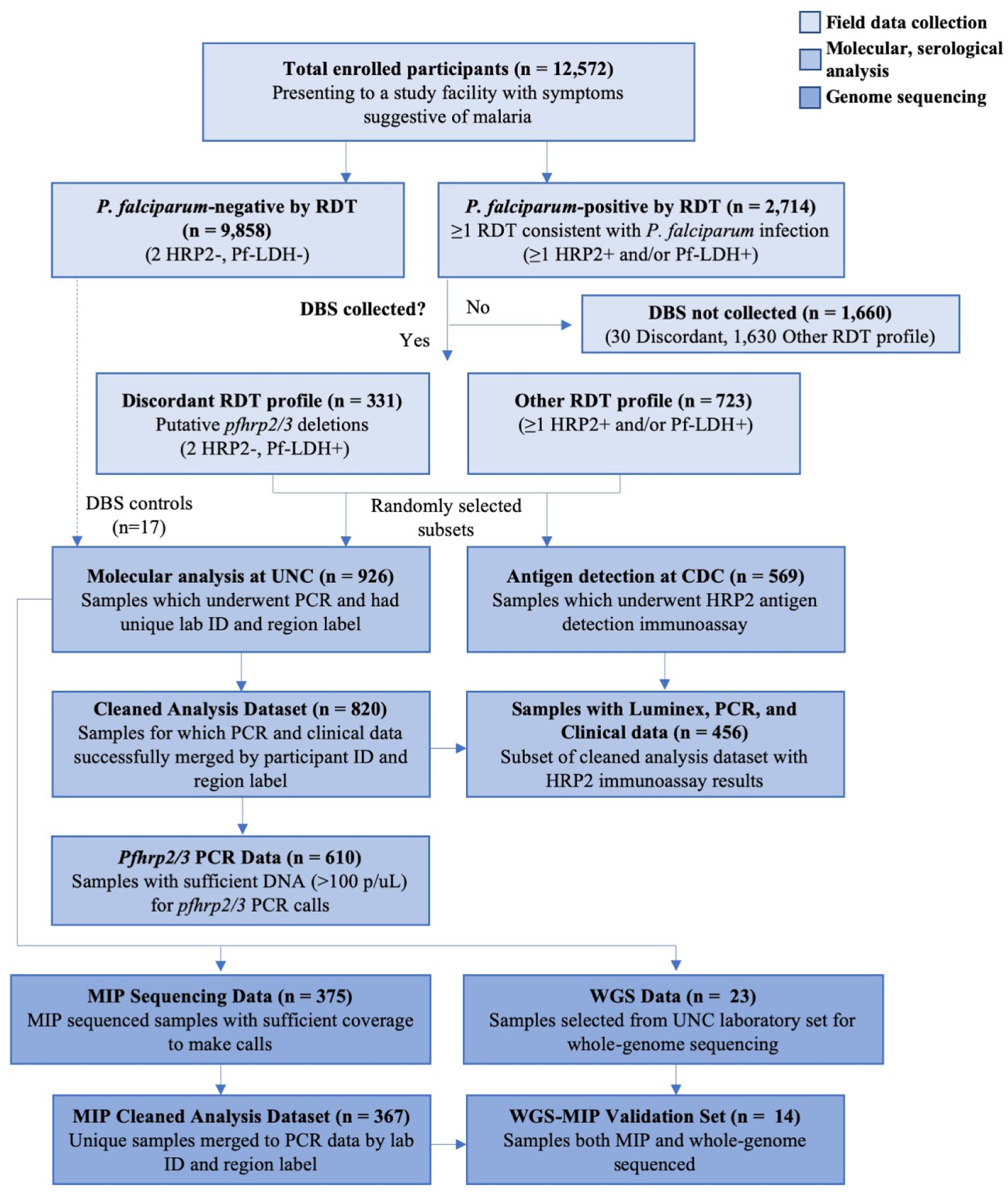
Study samples and assays performed.

**Supplementary Figure 2.**
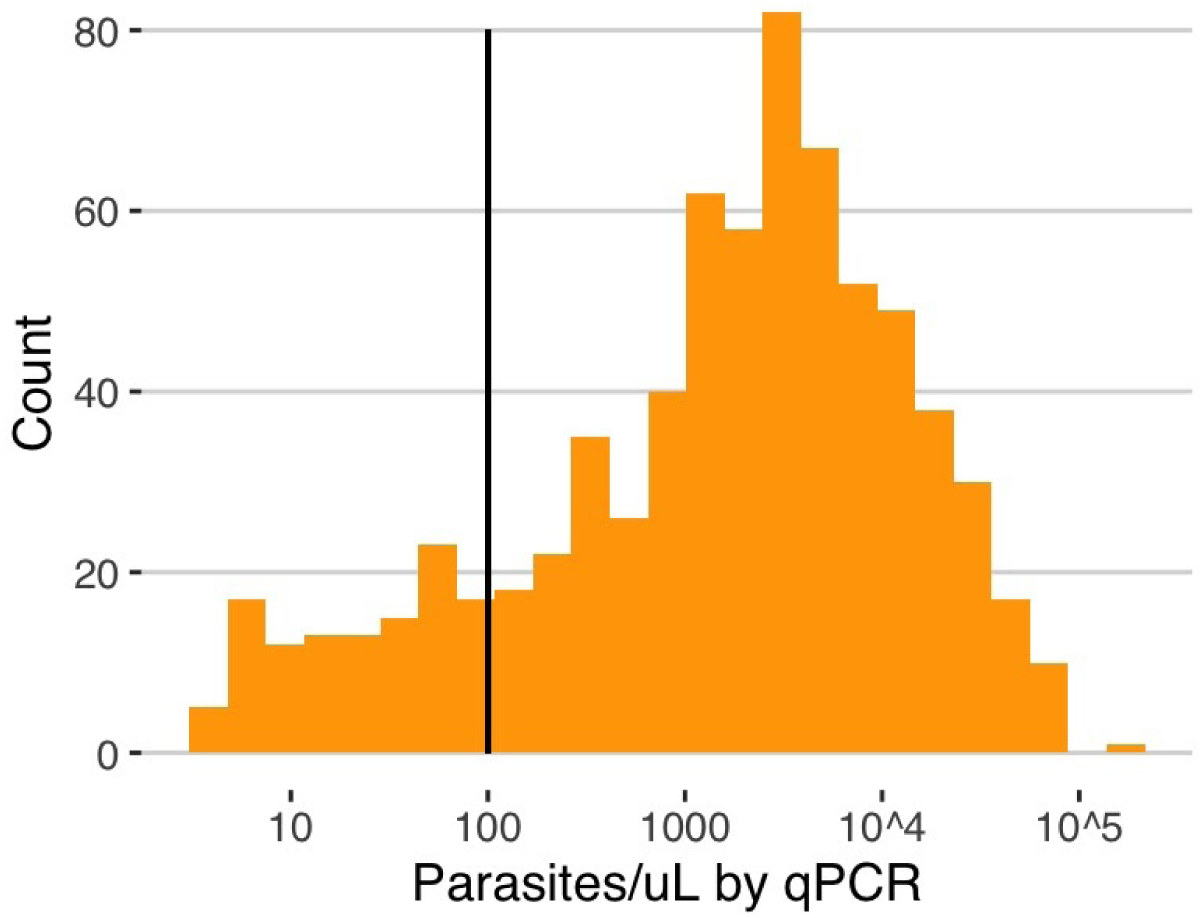
Parasite density distribution. *Pfldh* quantitative PCR (qPCR) results used to assess parasite density and determine which samples were eligible for *pfhrp2/3* deletion genotyping using a series of PCR assays. *Pfhrp2/3* deletions were only called in samples with >100 parasites/µL (solid line).

**Supplementary Figure 3.**
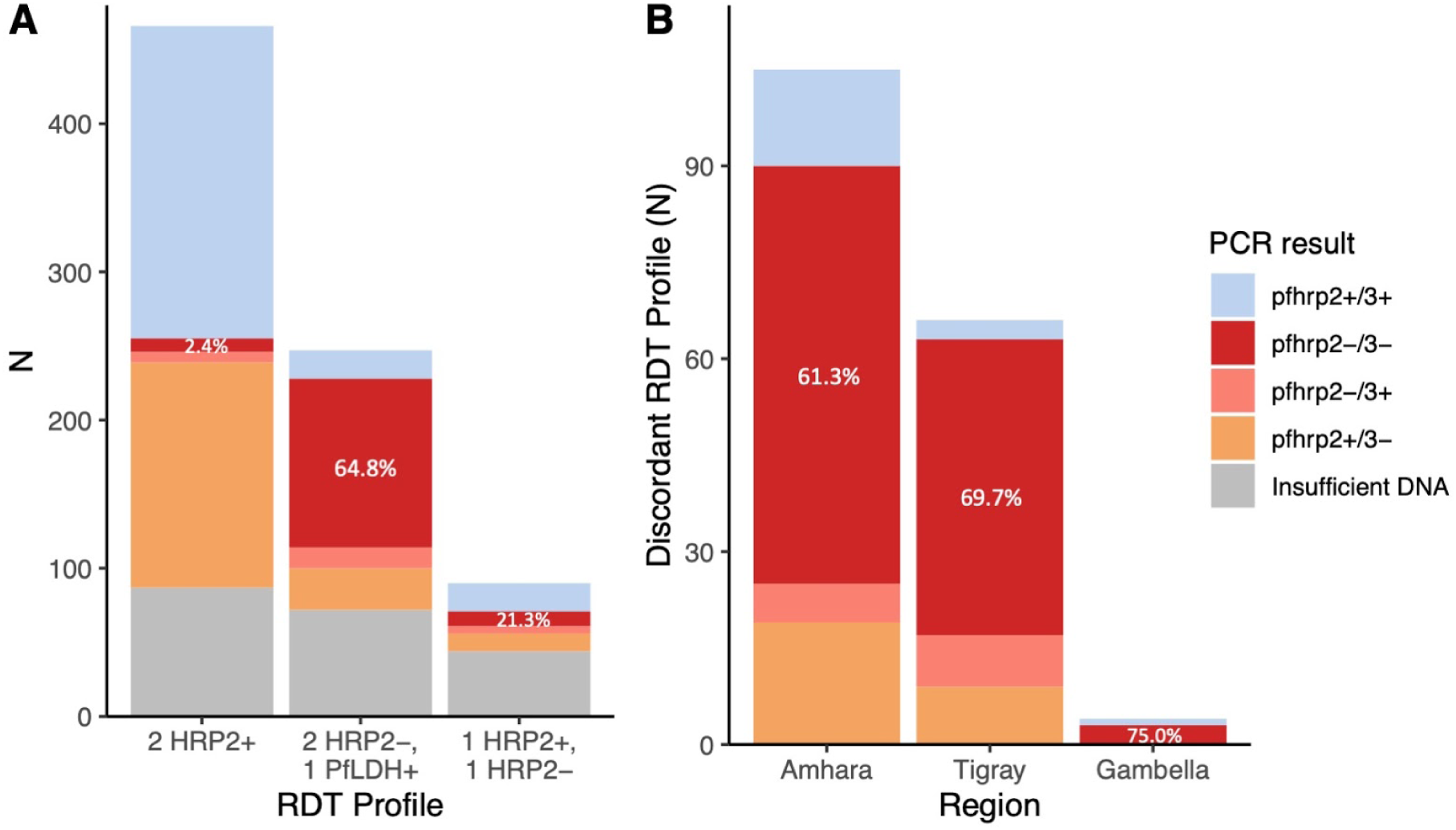
Pfhrp2 and pfhrp3 PCR results by RDT profile. A) Concordance between RDT profile and PCR *pfhrp2/3* result for *P. falciparum* samples with >100 parasites/µL. B) *Pfhrp2/3* PCR results for participants with the discordant RDT profile and sufficient DNA for molecular analysis (n = 176), by study region.

**Supplementary Figure 4.**
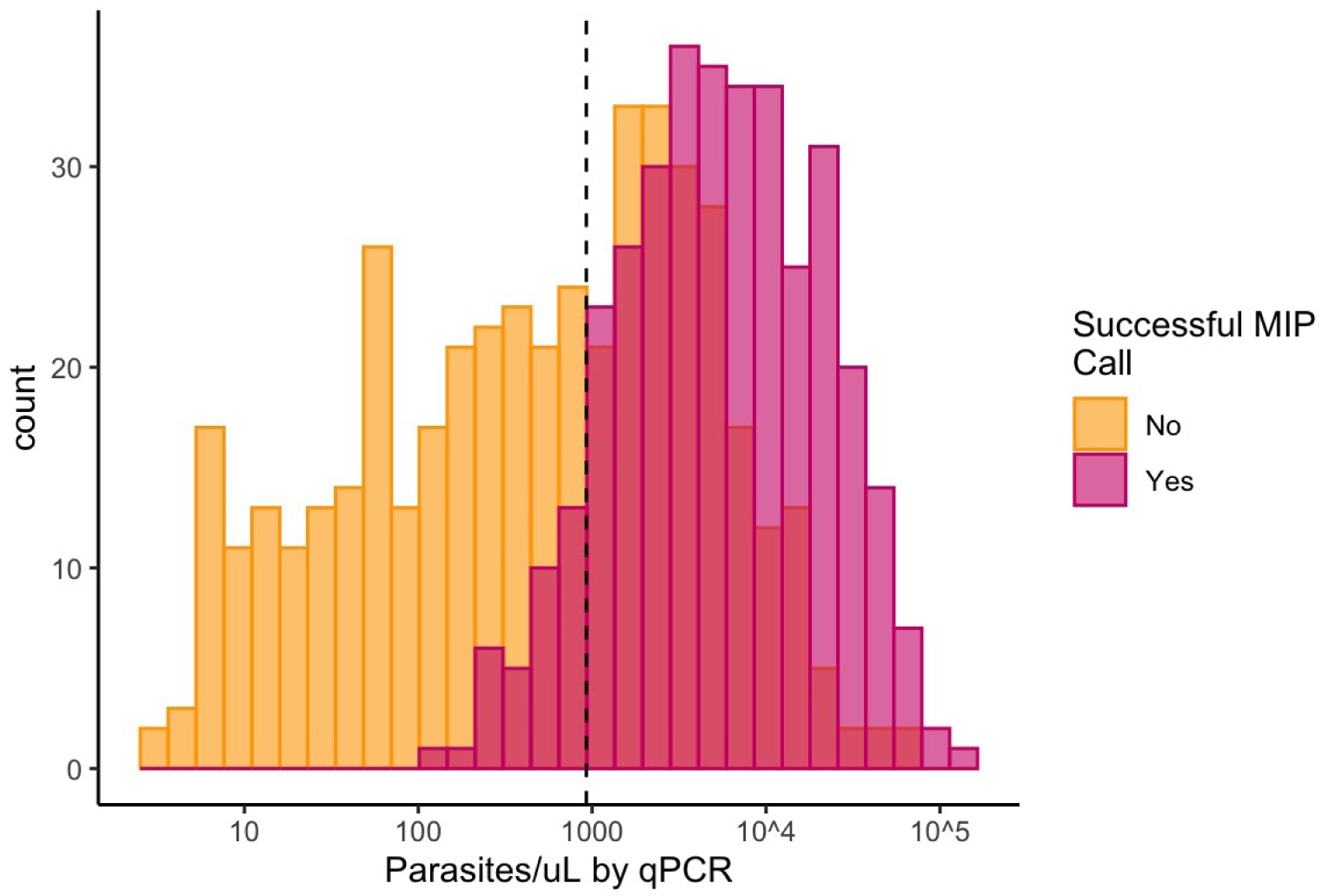
Successful MIP deletion calls versus qPCR parasite density. Comparison of MIP call results and qPCR parasite densities suggests a project-specific threshold for MIP calling of approximately 925 p/µL of whole blood (dashed line).

**Supplementary Figure 5.**
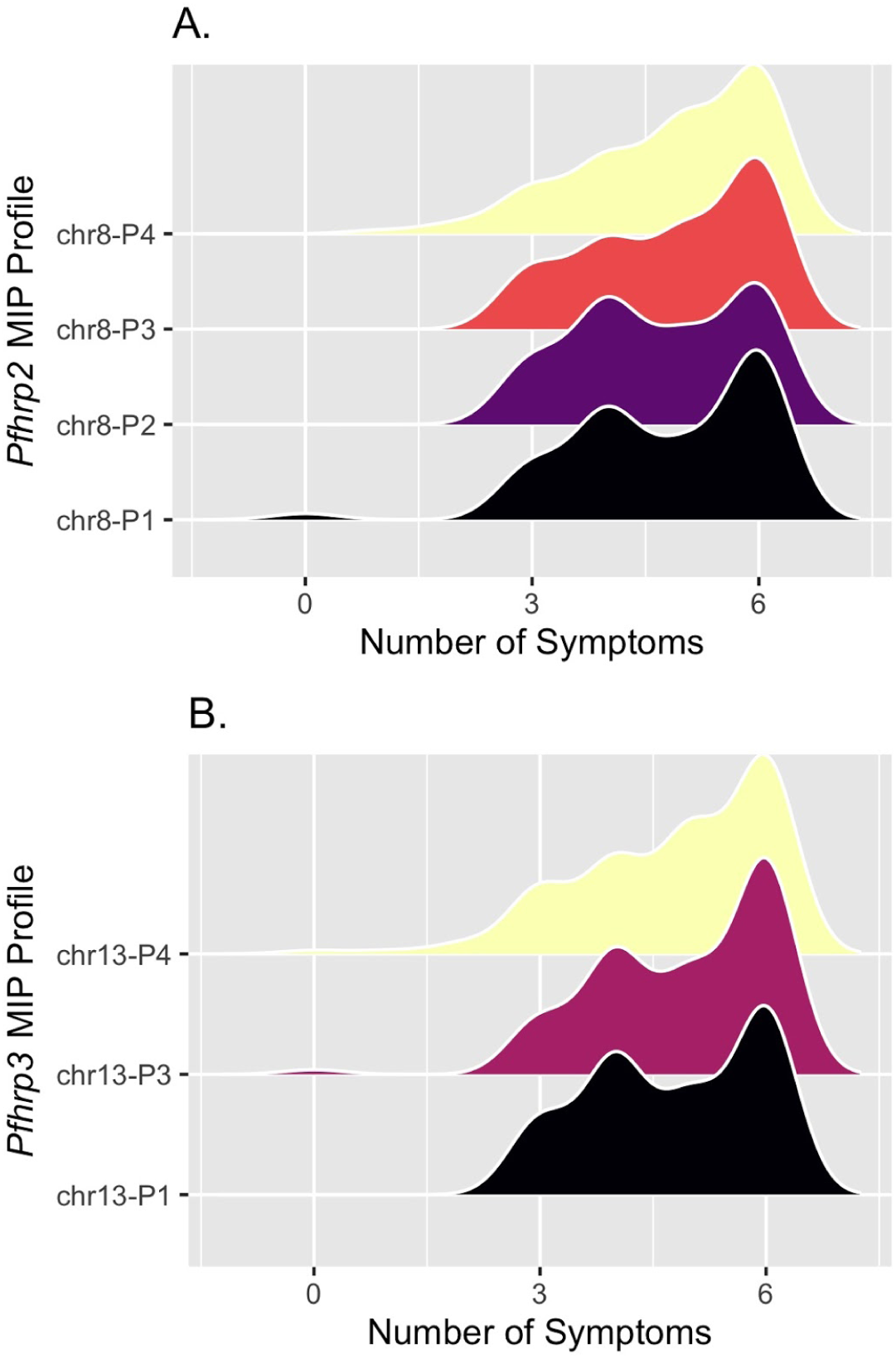
Disease severity by subtelomeric structural profile. Smoothed distribution of disease severity for each of the broader deletion breakpoint haplotypes along chromosomes 8 (A) and 13 (B) identified by MIP genomic enrichment. The total number of symptoms with which participants presented was used to estimate disease severity, with all participants evaluated for: fever, headache, joint pain, feeling cold, nausea, and loss of appetite. Profile chr13-P2 was excluded for its small sample size (n=1).

**Supplementary Figure 6.**
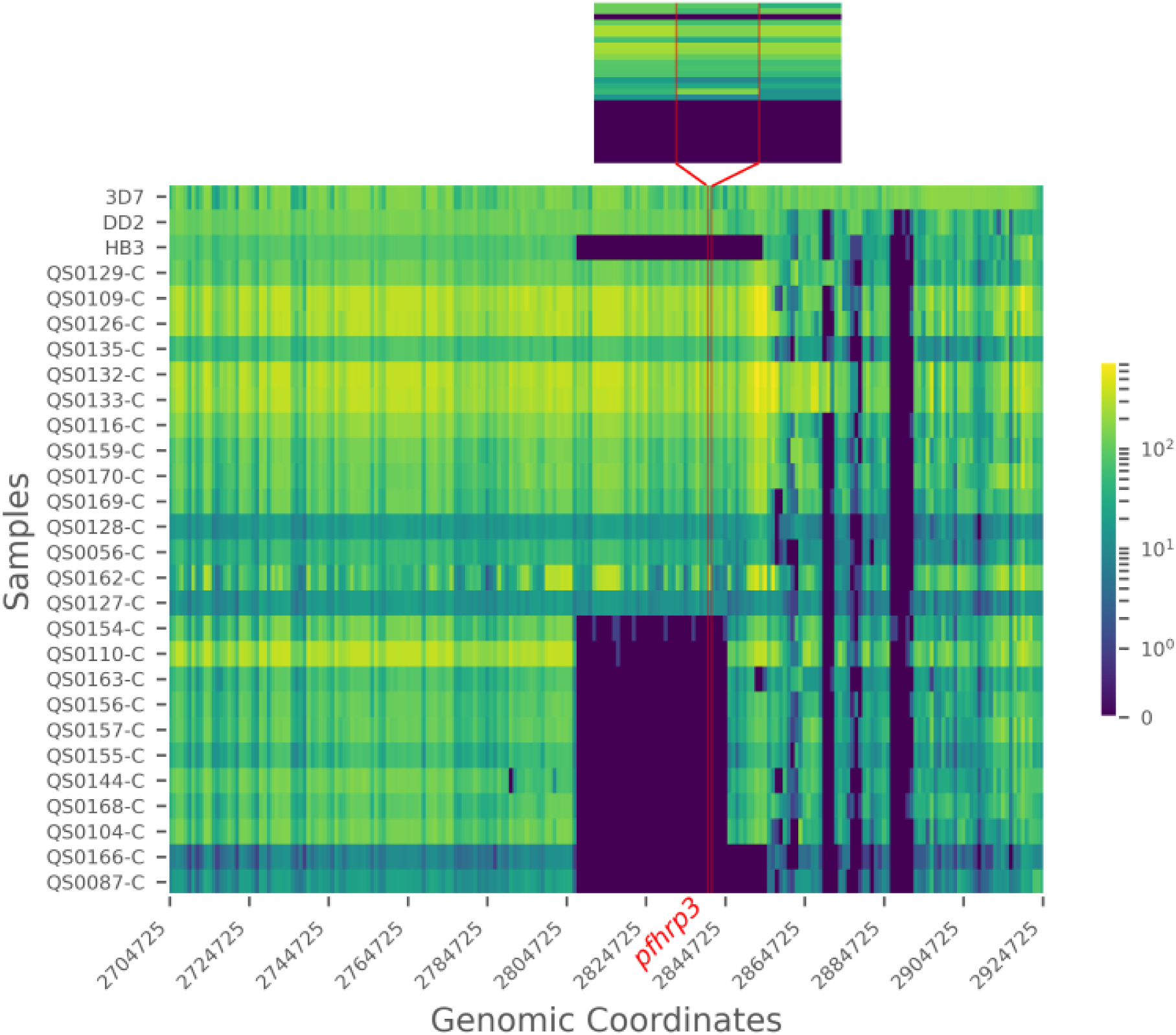
Chromosome 13 telomeric-end coverage (aligned reads/locus) plot of WGS control strains and 25 published Ethiopian genomes from 2013-2015 (MalariaGEN). The location of *pfhrp3* is indicated by vertical red lines. Inset showing *pfhrp3* and 1 kilobase flanking genomic region. Large subtelomeric deletions containing *pfhrp3* are apparent in the laboratory strain HB3, as well as 11 samples from Ethiopia.

**Supplementary Figure 7.**
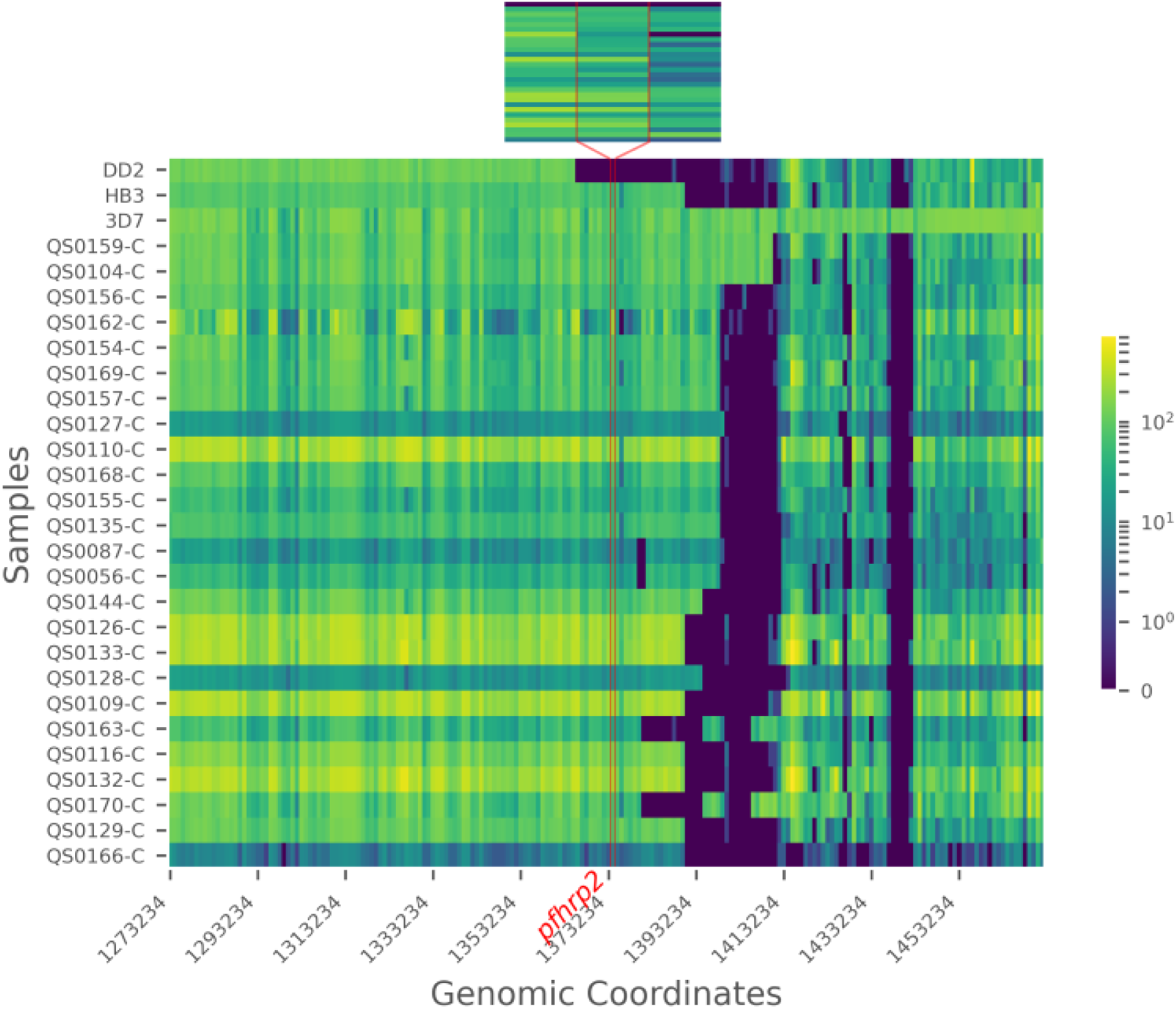
Chromosome 8 telomeric-end coverage (aligned reads/locus) plot of WGS control strains and 25 published Ethiopian genomes from 2013-2015 (MalariaGEN). The location of *pfhrp2* is indicated by vertical red lines. Inset showing *pfhrp2* and 1 kilobase flanking genomic region. Large subtelomeric deletions compared to the reference strain 3D7 are apparent in most samples. None of the deletions involve *pfhrp2* except the DD2 strain.

**Supplementary Figure 8.**
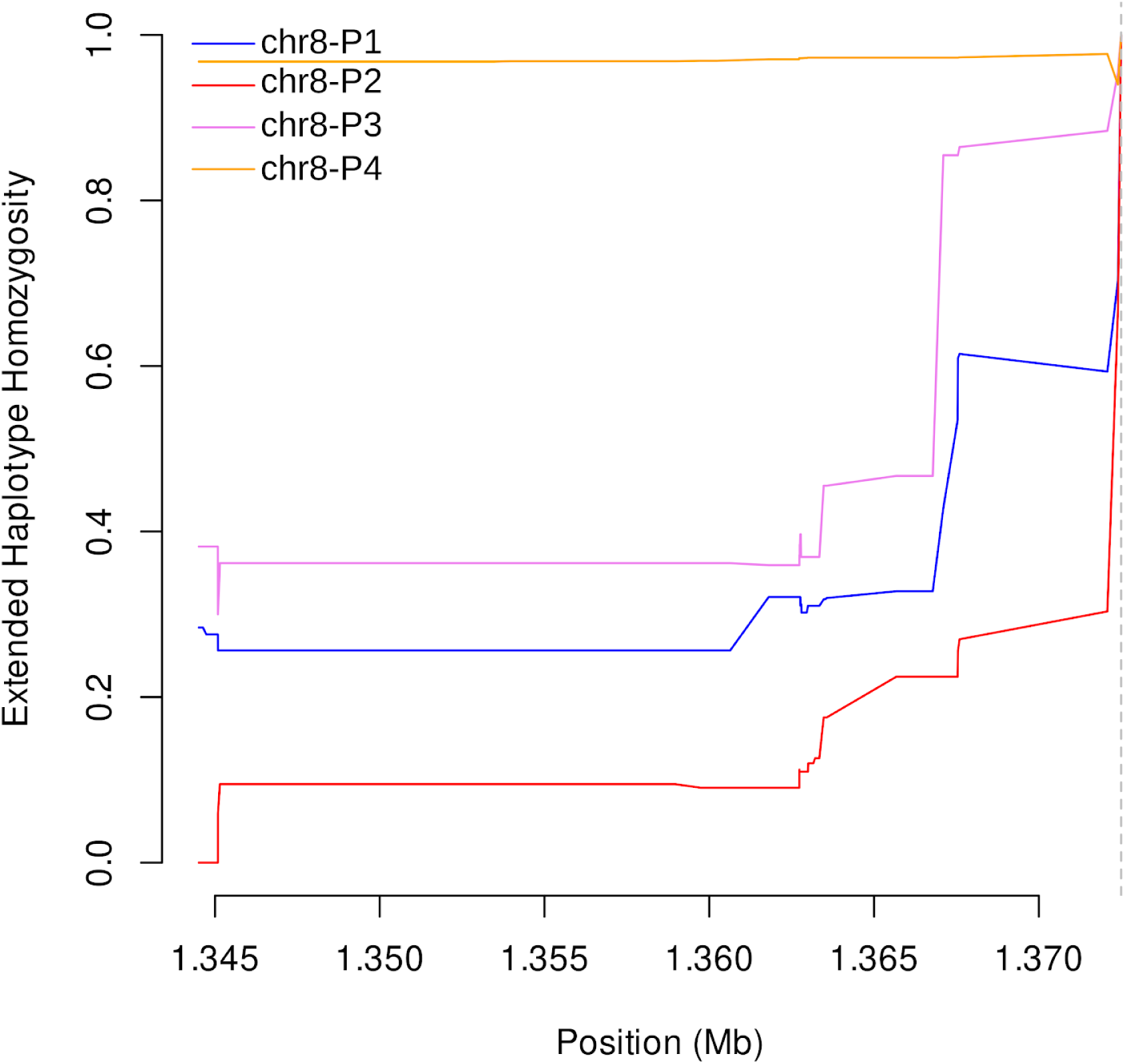
Extended haplotype homozygosity centromeric to chromosome 8 subtelomeric structural profiles P1-P4 using MIP data. The only *pfhrp2-*deleted profile (chr8-P4) showed sustained EHH, whereas EHH quickly broke down for the *pfhrp2*-intact profiles (P1-P3). A vertical dashed line on the right marks the centromeric end of the profile 4 (*pfhrp2)* deletion.

**Supplementary Figure 9.**
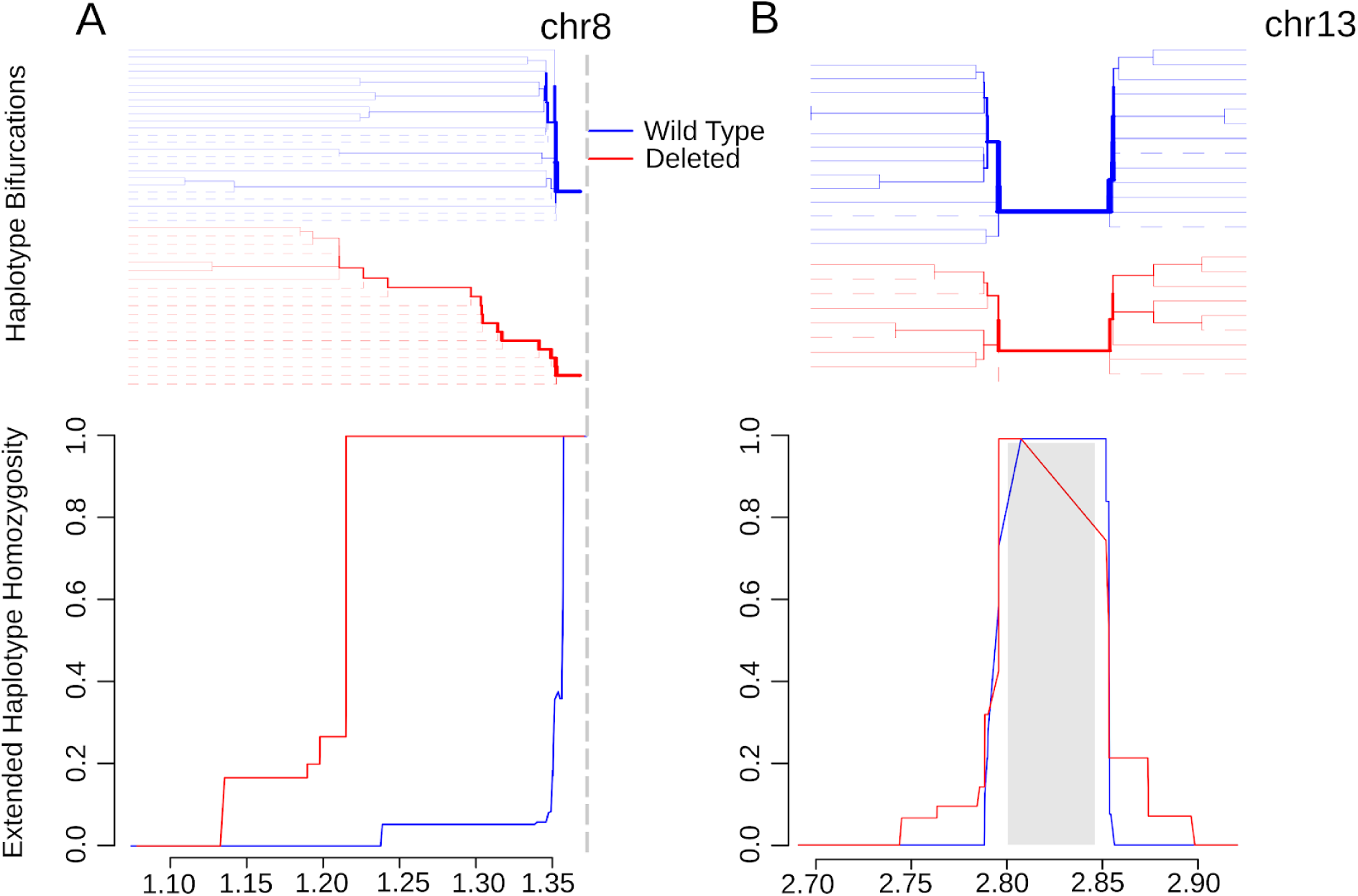
Extended haplotype homozygosity (bottom) and the bifurcation diagrams showing haplotype branching (top) centromeric to the pfhrp2 (A) and surrounding pfhrp3 (B) deletions based on WGS data. Vertical dashed line indicating the centromeric end of the chromosome 8 deletion (A). Gray box demarcating the chromosome 13 deletion (B). Abbreviations: Mb, mega-base.

**Supplementary Figure 10.**
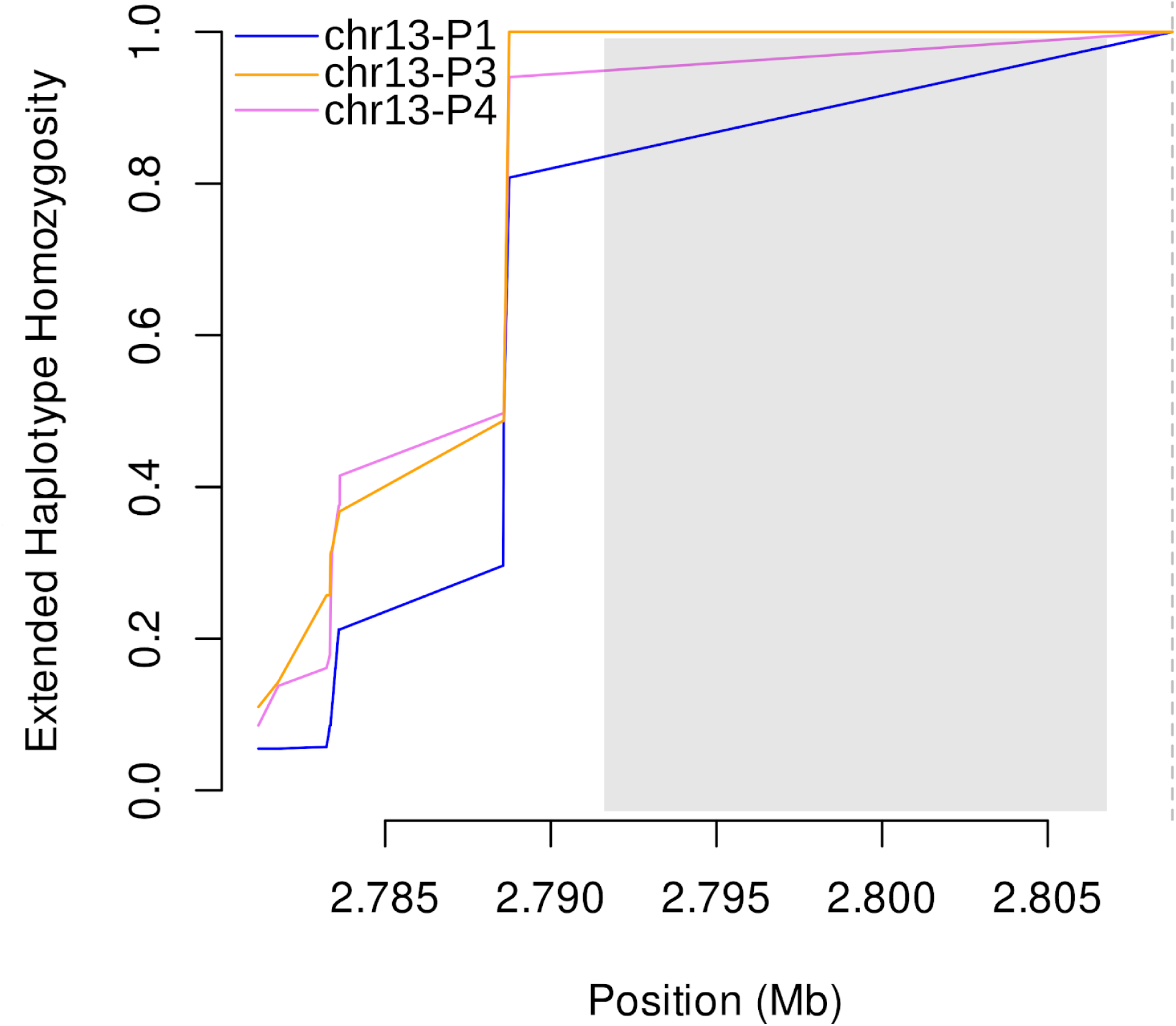
Extended haplotype homozygosity centromeric to chromosome 13 subtelomeric structural profiles P1, P3 and P4 using MIP data. Chr13-P2 profile was observed in only one sample and not included in the haplotype analysis. EHH quickly brown down for all profiles (P1: *pfhrp3-*intact, P3-P4: *pfhrp3-*deleted). A vertical dashed line on the right marks the centromeric end of the P3 and P4 deletions. No variants in the duplicated segment (gray box) were used in the EHH analysis.

